# Estimating the strength of symptom propagation from primary-secondary case pair data

**DOI:** 10.64898/2026.04.07.26350037

**Authors:** Phoebe Asplin, Rebecca Mancy, Matt J. Keeling, Edward M. Hill

## Abstract

Evidence indicates that infection from a symptomatic individual increases the risk of secondary cases developing symptoms, for a range of diseases. However, the magnitude of this effect has not previously been quantified. In this study, we use data on primary-secondary case pairs and their symptom status to quantify this increase in risk, which we refer to as the strength of symptom propagation.

We use publicly available data from the COVID-19 pandemic across three settings: households in England (*n* = 190) and Israel (*n* = 873), and contact tracing in Norway (*n* = 11, 192). We estimate the risk of being symptomatic as 14%, 12%, and 17% higher when infected by a symptomatic individual compared to being infected by an asymptomatic individual in the England, Israel, and Norway datasets, respectively (95% CIs: 0–72%, 3–23%, and 13–21%). These positive estimates remained after adjusting for age-dependent effects in the two datasets that provided age information (England households: 42% (1–75%), and Israel households: 11% (3–20%)).

We validated our methodology using synthetic data to assess accuracy and data requirements. We show that as few as 100 primary-secondary case pairs yield reasonable estimates of the strength of symptom propagation, while 1,000 pairs produce consistently low errors across replicates. Estimates were robust to severity-dependent reporting bias. The framework naturally extends to correct for age-related confounding.

Together, these results provide robust evidence for symptom propagation of SARS-CoV-2 and deliver consistent estimates of around 15% of its magnitude. Our approach requires minimal data and is readily applicable to a wide range of pathogens and epidemiological settings.

## 1 Introduction

Evidence suggests that symptom severity may be correlated between primary and secondary cases in transmission chains of respiratory pathogens, including SARS-CoV-2 and influenza. Such correlations can arise from epidemiological transmission mechanisms, such as the route of transmission or the dose of pathogen with which an individual is infected [1, 2]. We refer to this phenomenon as symptom propagation and have outlined evidence for these epidemiological mechanisms in our previous in-depth review [3].

Simply knowing whether symptom propagation occurs for a particular pathogen is valuable for public health interventions that can take advantage of this effect, for example when considering severity-dependent isolation strategies [3]. Furthermore, the impact of symptom propagation on epidemiological outcomes and the effectiveness of interventions can also depend on its strength [4]. We model symptom propagation through a single parameter, *α*, representing the strength of symptom propagation: a secondary case copies the symptom severity of the primary case with probability *α*; otherwise with probability 1 − *α*, the symptom severity of the secondary case is assigned randomly according to the baseline probability of having severe disease, *ν*. We align the parameter *ν* with the idea of ‘virulence’ – a measure of the intrinsic severity of symptoms generated by the pathogen. We can expand this model to include age groups by allowing *ν* to depend on the age of the secondary case.

Despite compelling biological evidence that symptom propagation occurs [3], there have been limited previous attempts to quantify its strength. Santermans *et al.* [5] used influenza population incidence data combined with social contact matrices to estimate the probability of symptomatic disease given infection by a symptomatic case or asymptomatic case, respectively. However, they found that they could only estimate these parameters if the relative infectiousness of symptomatic and asymptomatic cases was known. Methi and Madslien [6] used COVID-19 contact tracing data to show that infections generated by asymptomatic SARS-CoV-2 cases were more likely to be asymptomatic themselves. However, the study focused on hypothesis testing rather than inference, and as such did not consider the strength of symptom propagation.

The attempted estimation of symptom propagation parameters may have been limited to date because it requires data on primary-secondary case pairs and their respective symptom severities. This kind of data, for example collected through contact tracing, is intensive to collect. Consequently, there is typically a reduced volume of primary-secondary case pair data compared to other epidemiological data streams, such as population incidence estimates [7]. Primary-secondary case pair data may also suffer from reporting bias towards more severe cases, because cases typically self-select or are identified on the basis of symptoms [8]. This severity reporting bias is of particular importance for SARS-CoV-2, where a relatively large proportion of infected individuals are asymptomatic [9, 10].

A further potential complication when estimating the strength of symptom propagation is the number of factors that influence symptom severity, beyond those relating to the causal epidemiological mechanisms underpinning symptom propagation. This is exacerbated by social mixing which is typically assortative, meaning that individuals are more likely to interact with those who are similar to themselves leading to these factors being correlated between primary-secondary case pairs [11]. The correlation of these factors in turn may lead to correlations in symptom severity. We focus on a well-studied example of assortative mixing: age-dependent mixing. Age is known to affect symptom severity across many respiratory pathogens [12, 13] and to also be a strong driver of contact patterns [14, 15]. Specifically, individuals are more likely to be of similar age to their contacts, thus leading to a correlation in their symptom severity. Indeed, age structure has previously been discussed as an additional mechanism explaining how correlations in symptom severity could arise [16].

In this study, we develop a methodology to estimate the strength of symptom propagation from data on primary-secondary case pairs. Using the model introduced in Asplin *et al.* [4], we derive a likelihood for the symptom propagation parameters. To estimate our parameters using this likelihood requires only four summary statistics: the number of primary-secondary case pairs of each combination of two symptom presentation categories (e.g. mild or severe, asymptomatic or symptomatic). We first estimate these parameters from synthetic data, assessing the impact of different assumptions regarding data availability (i.e. the volume of data available for a given type of data) and the robustness of our results to biases in case reporting according to severity. We then extend our methodology to account for other factors, focusing on age-dependent effects as an example. Finally, we apply our methodology to estimate the strength of symptom propagation from three separate, country-specific SARS-CoV-2 infection and COVID-19 disease associated data sets.

We demonstrate that data on a relatively small number (100) of synthetic primary-secondary case pairs is sufficient to obtain a reasonable estimate of the strength of symptom propagation. Increasing the number of primary-secondary case pairs used in the analysis led to more accurate strength of symptom propagation estimates. Our estimates were robust to all reporting bias scenarios considered and a straightforward extension to our methodology effectively accounted for age-dependent effects. Applying our methodology to the real-world data sets from the COVID-19 pandemic, we found positive estimates for the strength of symptom propagation of SARS-CoV-2, indicating correlations in symptom severity between primary and secondary cases. Overall, we provide a practical tool for estimating the strength of symptom propagation that has minimal data requirements, with its generalisability enabling application across a wide range of pathogens and epidemiological settings.

## 2 Methods

We developed a methodology to estimate the strength of symptom propagation from statistics on the frequency of combinations of symptom presentations between primary-secondary case pairs. We used both synthetic data generated by an epidemiological model and real-world data. First, we describe the symptom propagation model and its parameterisation (Section 2.1). Second, we present our synthetic data generation and sampling methodology with and without age-dependence (Section 2.2). Third, we describe how we constructed the likelihood functions and computed parameter estimates for the symptom propagation parameters (Section 2.3). Finally, in Section 2.4, we provide details of how we apply our methodology to estimate the strength of symptom propagation from the three COVID-19 real-world datasets that we analyse in Section 3.2.

### 2.1 Defining the strength of symptom propagation

We considered a case in which symptom severity is of two types: mild or severe (e.g. asymptomatic versus symptomatic), which we denote using the subscripts *m* and *s*. We incorporated symptom severity and symptom propagation into the model framework through two key parameters, which we term the symptom propagation parameters: *α*, the strength of symptom propagation; and *ν*, the baseline probability of the pathogen causing severe disease in the absence of propagation effects. The parameter *ν* is aligned with the idea of ‘virulence’ – a measure of the intrinsic severity of symptoms generated by the pathogen.

In our epidemiological model, symptom propagation is modelled as follows. With probability *α*, an infected individual copies the symptom severity of their infector, whilst with probability 1 − *α*, their symptom severity is assigned randomly according to the underlying probability of severe disease, *ν* (as depicted in Fig. 1). When *α* = 0, the symptom severity of an infected individual has no dependence on the infector’s symptom severity; instead, the symptom severity of the infected individual depends entirely on *ν* – this corresponds to the typical assumption applied to compartmental infectious disease models that take severity into account. When *α* = 1, the symptom severity of an infected individual is wholly dependent on that of their infector, meaning that symptom severity is always passed on with infection. A parameterisation of *α* = 1 is akin to a two-strain model, in which one strain causes mild symptoms only, and the other strain causes severe symptoms only. Full details and analysis of this model are available in Asplin *et al.* [4].

**Fig. 1.**
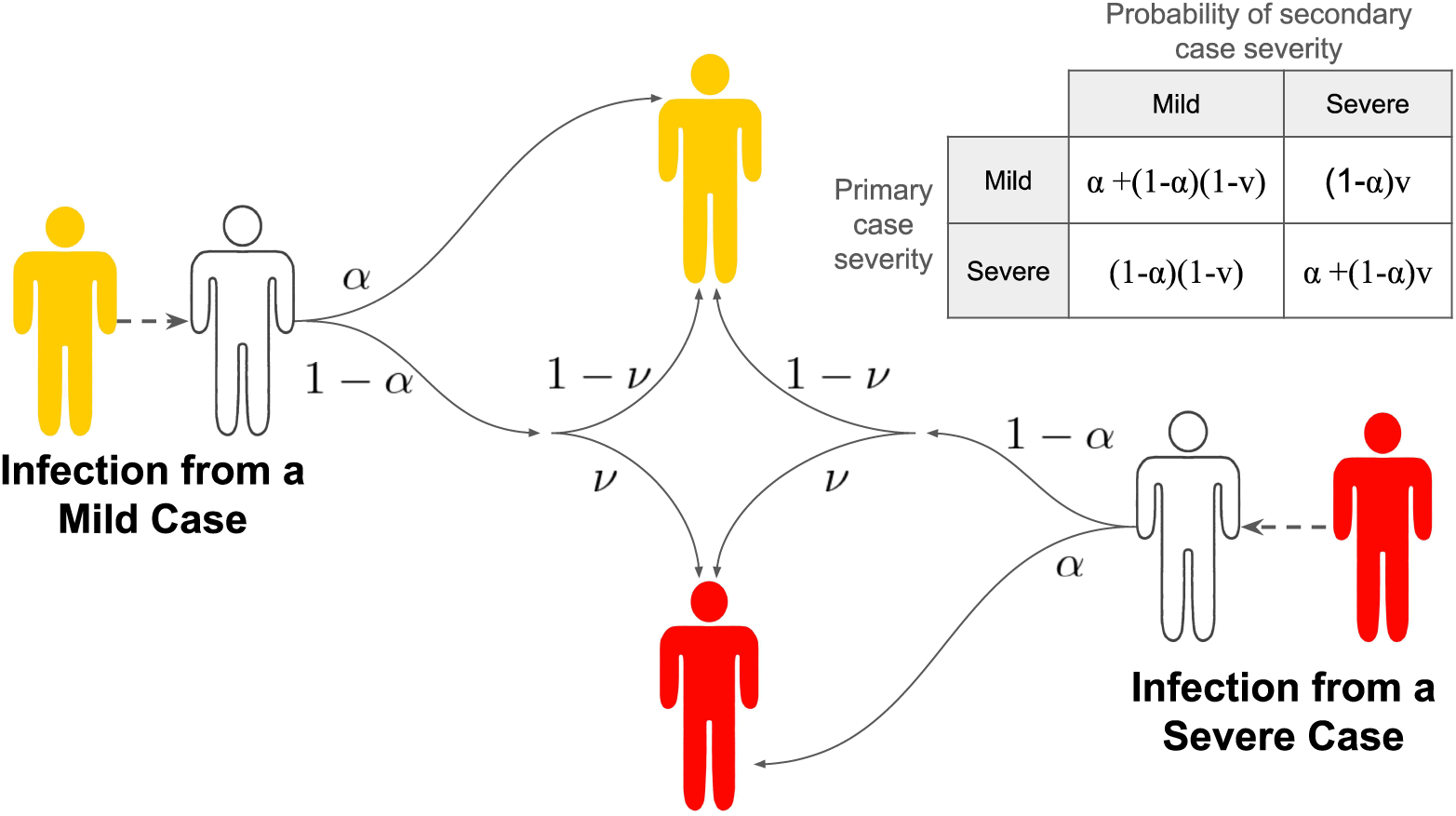
Schematic showing how symptom severity is determined in the symptom propagation model according to the two parameters characterising symptom propagation, *α* and *ν*. White-shaded individuals correspond to those susceptible to infection, yellow-shaded individuals correspond to infectious cases with mild symptoms, and red-shaded individuals correspond to infectious cases with severe symptoms. The values on the arrows show the corresponding probability. An infected individual has probability *α* of copying the symptom severity of their infector and a probability 1 − *α* of reverting to the baseline probability of having severe disease, i.e. they develop severe disease with probability *ν*.

### 2.2 Generating synthetic data

Using a compartmental model where individuals are either susceptible, infected or recovered, we simulated an epidemic over time, tracking the links between primary and secondary cases and their symptom severity. There are two versions of the model. First, in the baseline model, individuals are assumed to mix randomly and all have the same baseline probability of severe disease *ν*; this model was used to generate age-free synthetic data. Second, in the age-structured model, there is age-dependent mixing and the baseline probability of severe disease is also age-dependent, giving rise to a set of values *ν_i_*; this model was used to generate age-dependent synthetic data. We simulated epidemics for varying ‘true’ strengths of symptom propagation and then randomly sampled synthetic data sets containing varying numbers of primary-secondary case pairs and under various reporting bias scenarios.

Here, we describe in turn: the parameters used to generate the age-free synthetic data (Section 2.2.1); the parameters used to generate the age-dependent synthetic data (Section 2.2.2); the algorithm for generating the synthetic data using the *τ*-leap approximation to the Gillespie algorithm, generating both age-free and age-dependent synthetic data (Section 2.2.3); how we sampled the synthetic data with and without reporting bias (Section 2.2.4).

#### 2.2.1 Age-free parameters

We generated synthetic case data for a range of values of the strength of symptom propagation, *α*. To allow us to focus on the effect of the magnitude of *α* on our ability to estimate *α* and *ν* from the synthetic data, we held constant the overall proportion of cases that were severe, denoted P = 0.5; this proportion combines cases that were severe due to symptom propagation and those that were severe due to random effects driven by the baseline probability of severe symptoms *ν*. Details of how P was held constant are provided in Supporting Information Section S1.1. We also fixed the value of the reproductive ratio at *R*_0_ = 3, in line with estimates for pandemic influenza and SARS-CoV-2 [17–20]. We achieved this by calibrating the transmission parameter *β_m_* associated with mild cases (as described in detail in the Supporting Information of Asplin *et al.* [4]).

We list in Table 1 the values of *R*_0_, P and other epidemiological parameters (transmission rates *β_m_* and *β_s_* and recovery rate *γ*) used to generate the synthetic data. We assumed the recovery rates for mild and severe cases were equal (*γ* = 1*/*5, equivalent to an average infectious period of 5 days), as was required for the methodology used to fix the overall proportion of cases that were severe, P (described in Supporting Information Section S1.1).

**Table 1.** Epidemiological parameter values for age-free data. In each parameterisation, we calibrated *β_m_* to maintain *R*_0_ = 3. All rates are in units of ‘per day’ (day*^−^*^1^), with the exception of P and *R*_0_, which are dimensionless.

| Parameter | Description | Value | Source |
| --- | --- | --- | --- |
| $R_0$ | Basic reproduction number | 3 | [17–20] |
| $P$ | Proportion of infections that are severe | $1/2$ | Assumed |
| $\beta_m$ | Mild transmission rate | $\beta_m$ | Calibrated value |
| $\beta_s$ | Severe transmission rate | $2\beta_m$ | Couch *et al.* [21] |
| $\gamma$ | Recovery rate | $1/5$ | Byrne *et al.* [22] Byrne *et al.* [22] |

#### 2.2.2 Age-dependent parameters

In our age-dependent analysis, we incorporated age-dependent effects in relation to both social mixing and symptom severity. Consequently, *ν* varied with the age of the individual being infected. With the increased model complexity it was not possible to keep the proportion of cases that were severe constant. We instead fixed the age-dependent *ν* values to be constant across values of *α*. As a result, we no longer fixed *R*_0_ and instead used a single fixed value of *β* = 0.25; this meant that *R*_0_ ranged between 1.25 and 2.5, depending on the value of *α*.

We based our age-dependent parameterisation on three age groups: 0–17 year olds, 18–64 year olds and 65+ year olds. We chose these age groups as they are commonly used within the literature to approximately separate between school-age, working-age and retired individuals. We assumed 20% of individuals were in the 0–17 age group, 60% 18–64 and 20% 65+, based on 2023 ONS estimates for England and Wales [23]. To investigate a scenario in which severity was highly age-dependent, we chose the *ν* values to be 0.1, 0.5 and 0.9 (for the 0–17 year old, 18–64 year old and 65+ year old age groups, respectively). To show the robustness of our results to our parameterisation, we provide in Supporting Information Section S4.2.1 analysis for an alternative model parameterisation using less extreme age-dependent *ν* values (*ν* = [0.2, 0.4, 0.6]) which are rounded down estimates for the proportion of individuals in each age group experiencing shortness of breath reported by Andrew *et al.* [24] for influenza-like illness.

In order to simulate realistic social mixing, we used a social contact matrix that determined the rate at which individuals mix with those from the same age class versus the other age classes. The rates were based on the POLYMOD social mixing survey, using data for the UK from 2008 [14]. We used the R package *socialmixr* to aggregate the values into our desired age groups.

We explored the robustness of our results to our parameter choices in the Supporting Information Section S4.2. First, when considering the robustness of our age-free results, we varied one of the following from the default parameter value, with all other model parameters remaining unchanged: the relative transmissibility of severe cases (*b* = 0.5, 1, 4), the proportion of cases that were severe (P = 0.2, 0.8), and different values of the reproductive ratio *R*_0_ (= 1.5, 10). We also considered the effect of stronger reporting bias: specifically, we increased the probability of reporting severe cases from two (the baseline) to four times as likely as mild cases. We generally observed similar results to our main analysis using default values for all model parameters.

We also considered the effect of varying the transmission rates for the age-dependent analysis from a baseline scenario in which the transmission rate of severe cases *β_s_* was twice that of mild cases to one in which it was four times higher, and then one in which the rate was the same, and one in which the transmission rate of severe cases was half that of mild cases. In the latter two situations, we increased *β_m_* from 0.25 to 0.5 to ensure that *R*_0_ remained greater than one. Finally, we considered the situation in which the transmission rate for severe cases was twice that of mild cases, but increased the overall transmissibility by adjusting *β_m_* to be 0.5. Our findings were consistent with the main text results.

#### 2.2.3 Algorithm for generating synthetic data

To generate the synthetic data, we ran simulated epidemics using stochastic simulation. For both the age-free and age-structured symptom propagation models, we used the *τ*-leap approximation for the Gillespie algorithm. Here we give an overview of the synthetic data generation procedure, and the full details of the algorithms used are provided in Supporting Information Section S1.2 and S1.3.

We assumed an overall population size of *N* = 100, 000 individuals (of which 20,000 were aged 0–17, 60,000 were aged 18–64 and 20,000 were 65+ when age-dependent data was generated). We chose this population size to balance two goals: ensuring there were enough cases so that samples drawn in each replicate could be considered independent, whilst keeping the population size small enough that assuming everyone could potentially interact remained realistic (as is assumed in the Gillespie algorithm). We assumed the outbreak started with one mild and one severe case in each age group and that all other individuals were susceptible. We generated a single synthetic dataset for each value of *α*, ranging from 0 to 0.7 in increments of 0.1.

We used a standard two-severity SIR compartmental model where individuals are categorised as either susceptible, infected (with mild or severe symptoms) or recovered. Infected individuals infect susceptible individuals at a rate *β_m_* or *β_s_*, depending on their symptom severity. When age-structure was included, this transmission rate was augmented by the social contact matrix such that individuals were more likely to infect those of similar ages to themselves. When a susceptible individual is infected, their symptom severity is then determined by their infector’s symptom severity, *α* and *ν* (as described in Section 2.1). When age-structure was included, *ν* depended on the infected individual’s age group. Infected individuals then recover at rate *γ*.

For each transmission event, we considered whether the primary and secondary case were each mild or severe, and counted up the number of pairs with each combination of severity levels. The age-free dataset thus ultimately consisted of the four statistics *C^m→m^, C^m→s^, C^s→m^, C^s→s^*, the total numbers of each type of infection event (i.e. counts of events of mild case generating a mild case, mild case generating a severe case, severe case generating a mild case, severe case generating a severe case).

For the age-dependent analysis, we stratified the outputs by age group, *i*, of the secondary case, such that we obtained four statistics per age group: 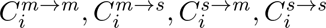. For example, 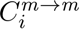 captures the number of transmission events from a mild primary case to a mild secondary case in age group *i*. Note that we do not require knowledge of the age group of the primary case, since this does not directly influence the severity of the secondary case; it only influences the age group of the secondary case, which we already have knowledge of.

#### 2.2.4 Sampling synthetic data

From our computational experiments, we wanted to understand how partial observation of infection events would affect our estimates for the strength of symptom propagation, *α*, since real-world data is likely to only represent a small sample of cases. To accomplish that aim, we varied the number of primary-secondary case pairs, *n*, we sampled from the overall synthetic data set (a sample of *C^m→m^, C^m→s^, C^s→m^, C^s→s^* in the age-free scenario). We sampled cases independently of when they occurred throughout the epidemic – small *n* can be thought of as corresponding to a low sampling intensity (rather than sampling occurring for a short period of time). We denote the sampled synthetic data set using ∼ notation, e.g. *C̃^m→m^* is a sample of *C^m→m^*. Therefore, *n* = *C̃^m→m^* + *C̃^m→s^* + *C̃^s→m^* + *C̃^s→s^* in the age-free scenario.

We then reasoned that mild cases in real-world data would be subject to under-reporting. We were interested in how this reporting bias would impact our parameter estimates. We considered four reporting bias scenarios. Each reporting scenario determines the relative probability of a case being sampled according to its symptom severity: (i) no reporting bias (default reporting scenario) – we sampled primary-secondary case pairs uniformly randomly from the synthetic data set; (ii) primary case reporting bias – primary severe cases were twice as likely to be detected as primary mild cases, and secondary cases would be equally likely to be detected regardless of severity (e.g. due to strict test-and-trace measures); (iii) secondary case reporting bias – primary cases would be equally likely to be detected regardless of severity and secondary severe cases were twice as likely to be detected as secondary mild cases (e.g. due to strict population-level testing but limited testing of contacts); (iv) primary-secondary case reporting bias – both primary and secondary cases were twice as likely to be detected if they were severe compared to mild cases. Under the reporting bias scenarios, compared to if both the primary and secondary cases were mild, a pair was twice as likely to be sampled if one of the primary or secondary cases was a severe case and four times as likely to be sampled if both the primary and secondary cases were severe.

For each combination of values of *n*, reporting bias scenario, and true value of *α*, we ran 1000 replicates, i.e. per replicate, we drew a sample of size *n* without replacement from the generated dataset for that true value of *α*, according to the reporting bias assumption.

For the age-dependent data set, we again sampled *n* primary-secondary case pairs in total. We sampled data for each age group in proportion to the percentage of the population they consisted of. For example, for the first age group (0–17 years, who comprised 20% of the overall population), we sampled 0.2*n* pairs in total from 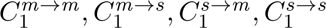. To restrict the complexity of the studied problem, we conducted these analyses assuming no reporting bias.

### 2.3 The likelihood of the symptom propagation parameters

We derived likelihoods from the sampled primary-secondary case pair data to allow us to obtain parameter estimates for *α* and *ν*. We first consider the age-free scenario, where we estimate *α* and a single value of *ν* from age-free data using maximum likelihood estimation. We then consider the age-dependent scenario where we estimate *α* and three age-dependent values of *ν* from age-structured data (a total of five parameters compared to only two for the age-free methodology). Calculating the likelihood over all possible combinations of these parameters to determine the maximum likelihood estimate would be very computationally intensive. We thus took a different approach to parameter inference, instead using Markov Chain Monte Carlo [25] rather than maximum likelihood estimation.

The likelihoods are stated below and the full derivations are provided in Supporting Information Section S1.4.

#### Age-free likelihood

We have a sample of aggregate contact tracing data, which describes the number of primary-secondary case pairs with each possible combination of symptom severities: *C̃m^→m^, C̃*^*m*^*^→s^, C̃*^*s*^*^→m^, C̃*^*s*^*^→s^*.

The model framework described in Section 2.1 specifies the probability that, given the primary case has severity *X*, the secondary case has severity *Y* (see Fig. 1). Given the data, the likelihood of *θ* = (*α, ν*) is given by

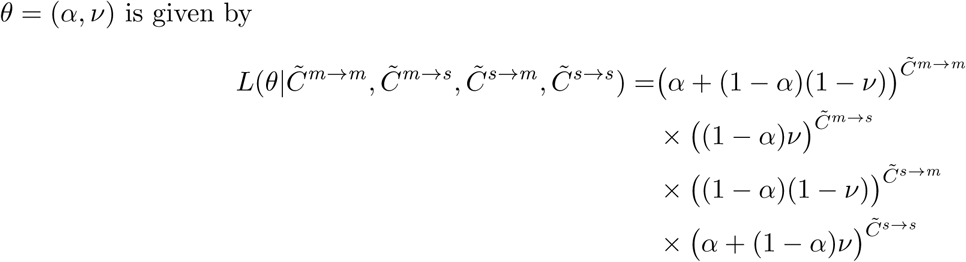

We denote the parameter values maximising this likelihood (the MLE) by *θ̂* = (*α̂, ν̂*).

#### Age-dependent likelihood

In the age-dependent setting, we assumed the availability of aggregate contract tracing data that is stratified by secondary case age group. We denote 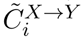 as the number of pairs with primary case severity X and secondary case severity Y, where the secondary case is in age group *i*. The likelihood of *θ* = (*α, ν*_1_*,…, ν_A_*), where *A* is the number of age groups, is given by

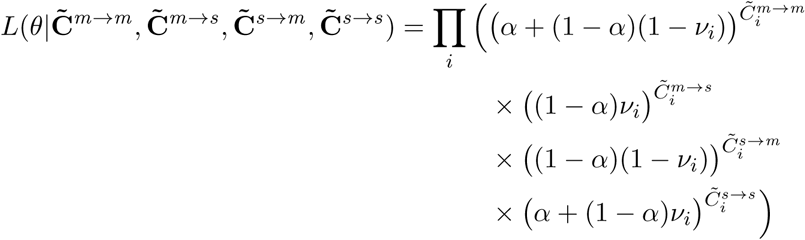

Our estimates *α̂* and *ν̂_i_* were now given by the means of the posterior distributions produced using MCMC, rather than the MLE. Specifically, we used the Adaptive Metropolis algorithm [26] with uniform priors, performing 30,000 iterations of which we discarded 3,000 as the burn-in period. For practical reasons associated with this change in methodology, we allowed *α* to become slightly negative to permit the estimate of *α* to be zero. Negative values of *α* are equivalent to mild cases being more likely to lead to severe cases and vice versa. Full details of how we applied MCMC are provided in Supporting Information Section S1.5.

### 2.4 Real-world data application: data from the COVID-19 pandemic

We applied our symptom propagation parameter estimation methodologies to three example real-world data sets from the COVID-19 pandemic: England household data [27], Israel household data [28] and Norway contact tracing data [6]. For the England and Israel household datasets, we computed the summary statistics from the individual-level data, using data on their household to construct links between cases. For the Norway contact tracing data we did not have access to the full dataset. We instead actively demonstrate that we can estimate symptom propagation parameters using solely publicly available summary statistics. The available data sets did not report symptom severity; however, they distinguished between asymptomatic and symptomatic cases. In our analysis, therefore, a mild case is one that is asymptomatic whereas a severe case is one that is symptomatic. This distinction is relevant for SARS-CoV-2 since a large proportion of cases are asymptomatic [29].

Although primary-secondary case pair data is directly collected through contact tracing, household data can also be used as a proxy. We construct primary-secondary case pair data under two key assumptions: (i) that additional infections within a household were likely a result of within-household transmission (supported by findings that between 70-85% of SARS-CoV-2 transmission occurs within households [30, 31]); (ii) that the individual who tested positive first was the primary case.

The summary statistics used for the parameter estimation for each of the three data sets is given in Table 2.

**Table 2.**
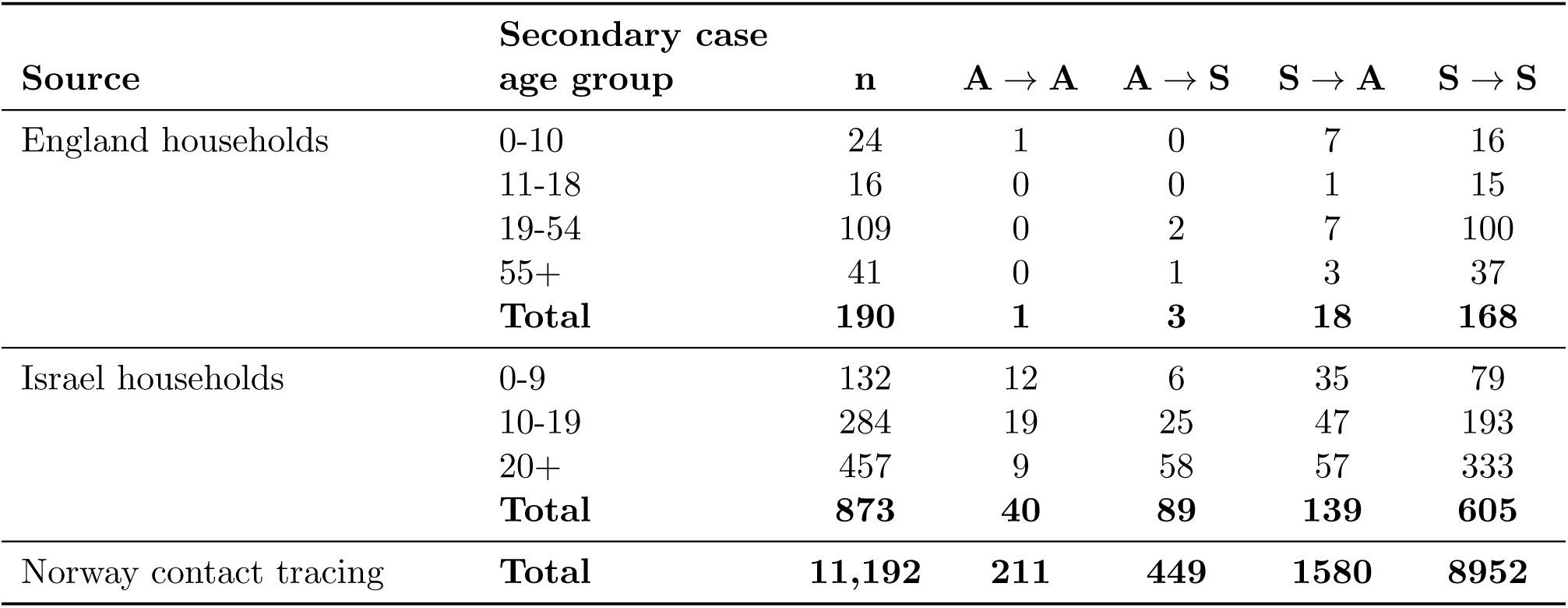
Summary case data used for parameter estimation stratified by secondary case age group. We provide the number of primary-secondary case pairs (*n*) from each study used in our parameter estimation, stratified by the age group of the secondary case, along with the symptom breakdown of all the pairs. Columns headed *X* → *Y* give the number of pairs where the primary case has symptom status *X* and the secondary case has symptom status *Y* (e.g. *A* → *A* is the number of pairs where both the primary and secondary cases were asymptomatic).

| Source | Secondary case | | $A \rightarrow A$ | $A \rightarrow S$ | $S \rightarrow A$ | $S \rightarrow S$ |
| --- | --- | --- | --- | --- | --- | --- |
| | age group | $n$ | | | | |
| England households | 0-10 | 24 | 1 | 0 | 7 | 16 |
|  | 11-18 | 16 | 0 | 0 | 1 | 15 |
|  | 19-54 | 109 | 0 | 2 | 7 | 100 |
|  | 55+ | 41 | 0 | 1 | 3 | 37 |
|  | **Total** | **190** | **1** | **3** | **18** | **168** |
| Israel households | 0-9 | 132 | 12 | 6 | 35 | 79 |
|  | 10-19 | 284 | 19 | 25 | 47 | 193 |
|  | 20+ | 457 | 9 | 58 | 57 | 333 |
|  | **Total** | **873** | **40** | **89** | **139** | **605** |
| Norway contact tracing | **Total** | **11,192** | **211** | **449** | **1580** | **8952** |

#### England household data

The first dataset is from a UKHSA household study in England conducted between February and August 2021 [27]. The study encompassed 227 households with a total of 559 participants. As such, there were a range of vaccination statuses and variants (full details in Table 3).

**Table 3.** Demographic details for real-world data sets. Details of the vaccination status, age group and variant for the England and Israel household data sets. Note that the Norway contact tracing data set is omitted here as we did not have access to this level of demographic information.

|  |  | England households |  | Israel households |  |
| --- | --- | --- | --- | --- | --- |
|  |  | Primary | Secondary | Primary | Secondary |
| **Vaccination status** | Not Vaccinated | 52 (27%) | 108 (57%) | 873 (100%) | 873 (100%) |
|  | Vaccinated | 138 (73%) | 82 (43%) | - | - |
|  | *One dose* | 67 (35%) | 38 (20%) | - | - |
|  | *Two doses* | 71 (37%) | 44 (23%) | - | - |
| **Age group** | 0-10 | 0 (0%) | 24 (13%) | - | - |
|  | 11-18 | 0 (0%) | 16 (8%) | - | - |
|  | 19-54 | 139 (73%) | 109 (57%) | - | - |
|  | 55+ | 51 (27%) | 41 (22%) | - | - |
|  | 0-9 | - | - | 95 (11%) | 132 (15%) |
|  | 10-19 | - | - | 161 (18%) | 284 (33%) |
|  | 20+ | - | - | 617 (71%) | 457 (52%) |
| **Variant** | Wildtype | 0 (0%) |  | 873 (100%) |  |
|  | Alpha | 112 (59%) |  | - |  |
|  | Delta | 78 (41%) |  | - |  |

In this study, index cases were identified through community testing. The index case and all consenting household members were interviewed to document symptom onset dates, which we used to determine if they were asymptomatic or symptomatic (with NA corresponding to asymptomatic). Household members were additionally tested via PCR at recruitment (day 1) and again on days 3 and 7. Full study details are available in the Supplementary Materials of Hart *et al.* [27].

We assumed the recorded ‘index case’ (identified from community testing) was the primary case and was responsible for infecting all secondary cases within the household. In Hart *et al.* [27], the authors assumed an individual was infected if they returned a positive PCR test at any point and/or developed at least one of the relevant symptoms. We replicated this approach in our analysis, assuming that individuals were infected if they tested positive and/or exhibited symptoms. This method meant that asymptomatic individuals were more likely to be omitted from the analysis because their inclusion depended on a positive PCR test on day 1, 3 or 7 after the primary case was recruited, which may have contributed to the small number of asymptomatic cases in the final data set (only four asymptomatic primary cases and nineteen asymptomatic secondary cases; see Table 2).

#### Israel household data

The second data set is from a household study in Bnei Brak, Israel [28]. This study took place between 17^th^ March and 3^rd^ May 2020. At that time, no individuals were vaccinated against SARS-CoV-2 and wild-type SARS-CoV-2 was the only variant in circulation. Because the goal of the study was to examine the role of children in transmission, the age groups recorded had a coarser resolution for adults; the recorded age groups were 0-9 years, 10-19 years and 20+ years, respectively (full details in Table 3).

In Bnei Brak, during the time period of the study, all members of a household in which a suspected case occurred were tested (full study details available in Dattner *et al.* [28]). Unlike the England data set, no individuals were marked as the index case. We assumed that the index case in each household was the individual who tested positive first. We assumed this individual was the primary case for all other infected individuals within their household. If multiple individuals tested positive on the same day, one was chosen as the primary case at random. Due to this requirement for a positive test to be considered a primary case, we subjectively chose to assume that only those with a positive test were infected, even if others reported symptoms. Even with this assumption, there were still many more symptomatic individuals than asymptomatic individuals in the dataset (694 symptomatic vs 179 asymptomatic secondary cases Table 2).

#### Norway contact tracing data

The third and final real-world data set we used was aggregate information from contact tracing from Oslo, Norway, collected between September 2020 and September 2021 [6]. This data set was not publicly available; however, the aggregate contact tracing data required for our analysis was described in Methi and Madslien [6]. Because of this, we did not have access to demographic information and the only data used in our analysis were the values listed in Table 2.

Vaccination statuses were recorded in the study but summary information was not publicly available. It was, however, stated that individuals had between 0 and 3 doses, and asymptomatic cases had a mean number of 0.07 doses, and symptomatic cases had a mean number of 0.12 doses. Similarly, age group data were not publicly available, but ages ranged from 0 to 105, with an asymptomatic case mean age of 24.1 and a symptomatic case mean age of 31.2. There was no discussion or record of variants; however, based on the time period (September 2020 – September 2021), likely circulating were a combination of wild-type, alpha, and delta variants [32].

Methi and Madslien [6] defined all positive cases identified through mass testing to be index cases. Close contacts were then identified through personal interviews with the index cases. They were then tested and symptom data was collected via telephone interview. Their analysis was restricted to only close contacts who tested positive within 14 days after the index case tested positive. Contacts with missing symptom information were also excluded. Finally, they excluded close contacts who were linked to multiple index cases if those index cases were not both asymptomatic or both symptomatic. The final count of included individuals was 11,192 positive close contacts from 7786 index cases (see Table 2 for symptom breakdown). Full study details are described in Methi and Madslien [6].

## 3 Results

Our analysis is formed of two main parts. Our main priority is estimating the strength of symptom propagation from real-world data, however we first need to verify the robustness of our methodology. Therefore, we first used synthetic data to analyse the data requirements of estimating the strength of symptom propagation, examining the impact of different assumptions regarding data availability, reporting bias according to case severity, and age-dependent effects (Section 3.1). Then, we applied our methodology to three example SARS-CoV-2 infection and COVID-19 disease data sets to estimate the strength of symptom propagation (Section 3.2).

### 3.1 Synthetic data results

In Asplin *et al.* [4], we show that the strength of symptom propagation impacts the effectiveness of vaccination strategies and even can affect the choice of interventions (e.g. whether transmission-blocking versus symptom attenuating vaccines are most effective). It is therefore important for public health professionals to understand whether symptom propagation is taking place, and further, to be able to accurately estimate the strength of symptom propagation.

Motivated by this, we examine three key outcome measures. First, we consider the uncertainty in our estimates: in any given replicate of our estimation procedure, how confident are we in our produced estimates. Second, we consider the accuracy of our estimates: across all 1,000 replicates, how close do we get to the true value on average. Finally, we consider the support for symptom propagation: across all of 1,000 replicates, how often can we be confident that symptom propagation is occurring (corresponding to our lower bound for *α* being greater than zero). This final outcome measure had two goals: to measure how often we failed to detect symptom propagation when it was occurring, and how often we found ‘false positives’ (support for symptom propagation when it was not occurring).

Specifically, we estimated the two parameters characterising symptom propagation, *α* and *ν*, from our synthetic primary-secondary case pair data. We examined how our assumptions regarding data availability, reporting bias and the true strength of symptom propagation impacted: (i) the uncertainty in inferred age-free estimates of *α* and *ν* (Section 3.1.1); (ii) the accuracy of the age-free estimates of *α* and the inferred support for symptom propagation, examining the effect of reporting bias on these results (Section 3.1.2); (iii) the accuracy of our age-dependent estimates and inferred support for symptom propagation from data with underlying age structure (Section 3.1.3).

#### 3.1.1 Uncertainty in symptom propagation parameters

In our initial scenario, we assumed there was no reporting bias associated with case severity. We explored how the 95% confidence regions (CRs) of our estimates *α̂* and *ν̂* varied with the number of pairs we had data for, *n*, and the ‘true’ strength of symptom propagation, *α*, used to generate the synthetic data (Fig. 2).

**Fig. 2.**
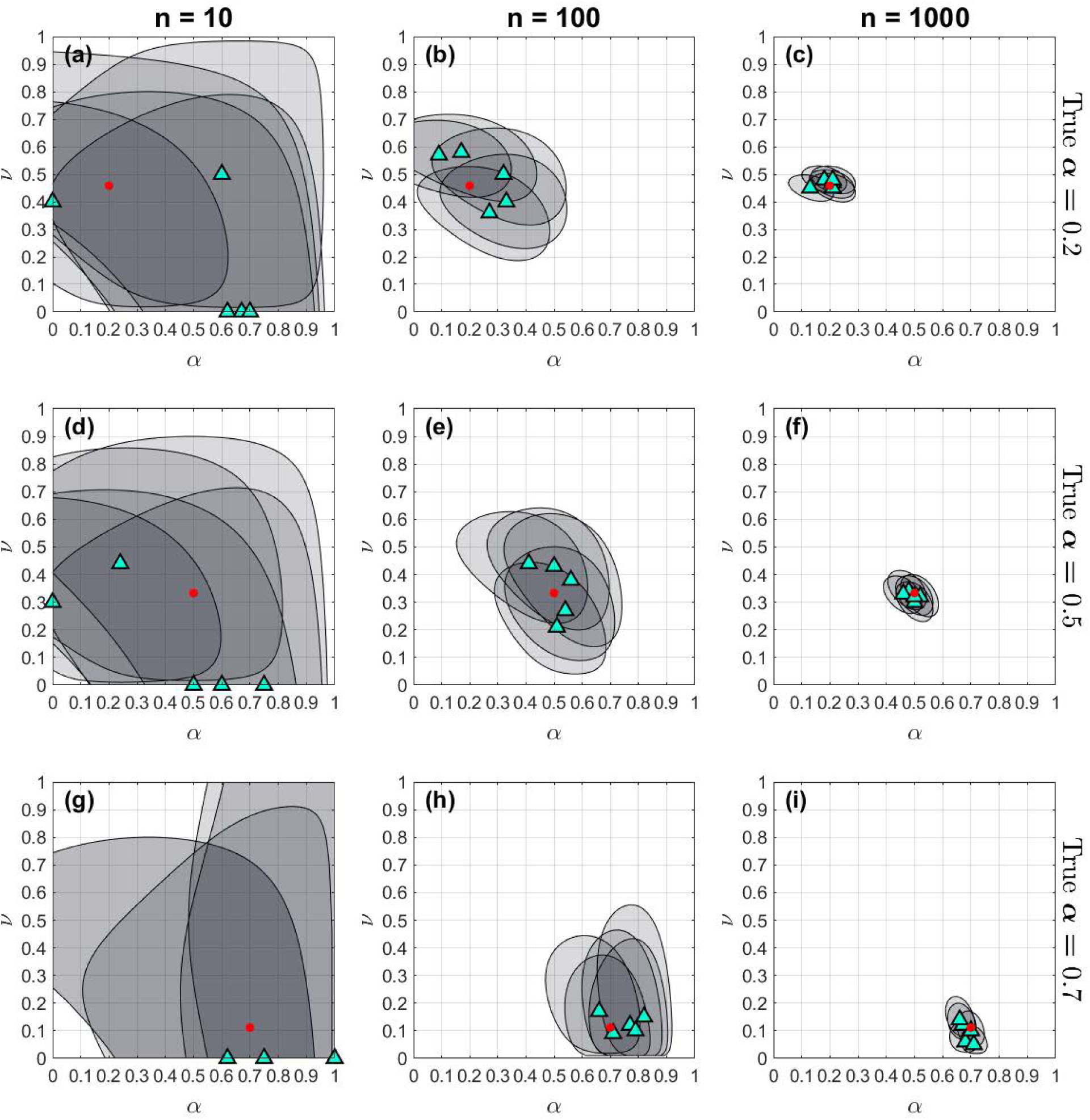
95% bivariate confidence regions for three values of *α* and three values of *n*. 95% confidence regions for the predicted value of *ν* and *α* for five replicates with varying value of *n* (the number of primary-secondary case pairs we had data for in each replicate): **(a, d, g)** 10, **(b, e, h)** 100, **(c, f, i)** 1,000. The cyan triangles indicate the MLE estimates for each replicate. The red dots correspond to the true values of *α*. The true value of *α* is varied across the rows: **(a-c)** *α* = 0.2, **(d-f)** *α* = 0.5, **(g-i)** *α* = 0.7.

We found that synthetic data on a larger number of primary-secondary case pairs allowed us to be less uncertain in our estimates for *α* and *ν*. For *n* = 10 (Fig. 2, first column), the 95% CRs were large, often containing the majority of parameter combinations (average width in the *α* direction of 0.82). There was also variation in the shape of the confidence region between replicates. In comparison, when we had synthetic data on a larger number of pairs (Fig. 2, second and third columns), the CRs were smaller and more consistent in shape (average width of 0.37 for *n* = 100 and 0.12 for *n* = 1, 000).

Interestingly, we also found that our uncertainty in the strength of symptom propagation was reduced when symptom propagation was stronger in the underlying data set. Consistently, the uncertainty in *α* (given by the width of the CR) was greater for smaller true *α* than for larger true *α*. The opposite was true of the uncertainty in *ν* (given by the height of the CR). Specifically, for *n* = 100, the average width (in the *α* direction) of the 95% CR was 0.43 when true *α* = 0.2 (Fig. 2(b)) compared to 0.29 when true *α* = 0.7 (Fig. 2(h)). For *n* = 1, 000, the average width of the 95% CR was 0.15 when true *α* = 0.2 (Fig. 2(b)) compared to 0.088 when true *α* = 0.7 (Fig. 2(h)).

#### 3.1.2 Accuracy of estimates for the strength of symptom propagation

We next considered the accuracy of our estimates for the strength of symptom propagation, *α̂*, produced by our inference procedure (Fig. 3, top row). Specifically, we investigated how close the maximum likelihood estimates (MLE) were to the true value of *α* used to generate the synthetic data (Fig. 3, top row). We investigate both the median error in *α* (shown by the solid markers) which indicates how accurate are we on average across all 1,000 replicates and a 95% uncertainty interval (shown by the translucent shaded regions, spanning the 2.5^th^ percentile to the 97.5^th^ percentile of the errors) which indicates the range of the errors seen across replicates. Therefore, a wide uncertainty interval indicates that more than 5% of replicates generated highly inaccurate estimates. Note that the solid grey shaded areas indicate infeasible errors, due to our estimates of *α* always being between 0 and 1 (e.g. for true *α* = 0, it is not possible for us to underestimate *α*).

**Fig. 3.**
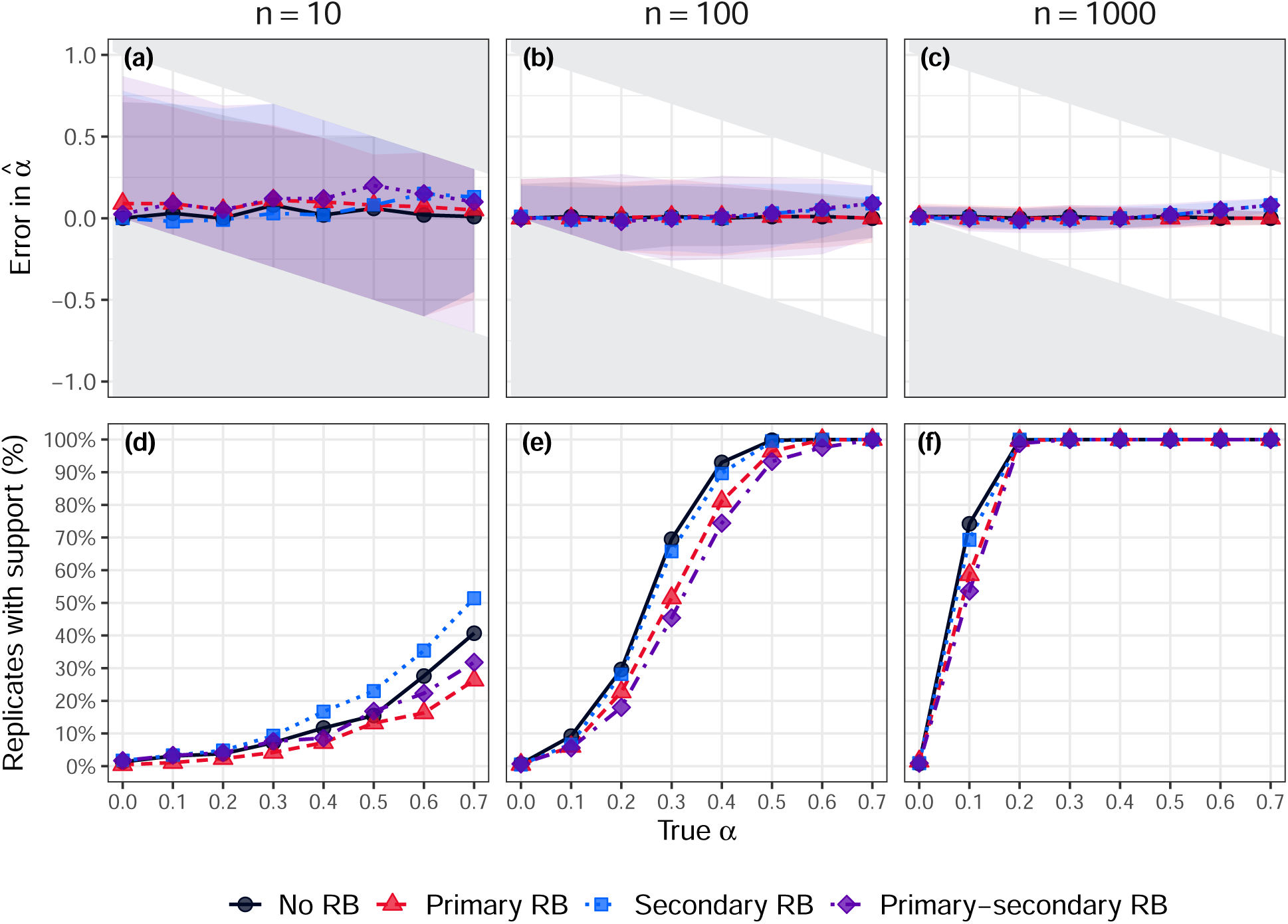
Error in the estimated values of *α̂* and the percentage of replicates with support for symptom propagation. We consider four reporting bias scenarios: no reporting bias (black circles), primary case reporting bias (red triangles), secondary case reporting bias (blue squares) and primary and secondary case reporting bias (purple diamonds). The top row shows the error in the estimates of *α̂*. A positive error corresponds to the estimated value being an overestimate of the true value of *α*. The solid grey regions correspond to infeasible errors, which are present due to the restriction of *α̂* being between 0 and 1. The points show the median errors and the lines show the interpolation between those points. The shaded area spans the 2.5th percentile to the 97.5th percentile (the 95% uncertainty interval). The bottom row gives the percentage of replicates (out of 1000) for which *α* = 0 was not within the 95% confidence region (i.e. support for symptom propagation). Across panels, we vary the assessed sample sizes *n* (i.e. the number of primary-secondary case pairs we had data for in each replicate): **(a, d)** 10, **(b, e)** 100, **(c, f)** 1,000.

Most notably, we found that the range of the errors was highly dependent the number of primary-secondary case pairs in the sampled synthetic data set: the maximum width of the uncertainty interval reduced by 63% as we increased from *n* = 10 to *n* = 100 and again by 64% as we increased from *n* = 100 to *n* = 1, 000. For *n* = 10 (Fig. 3(a)), the 95% uncertainty intervals were extremely broad, such that a single sample could return a highly erroneous estimate of *α*. Therefore, if we only had data on 10 primary-secondary case pairs, we could not be confident that our estimates would be accurate. In contrast, for larger *n* the amount to which we may over- or under-estimate *α* is greatly reduced (95% uncertainty intervals ranged between −0.26 and 0.27 for *n* = 100 and −0.09 to 0.12 for *n* = 1, 000).

When considering the no reporting bias case (black circles), the median errors in *α̂* were consistently close to zero across all values of *n*, and the uncertainty intervals were symmetrical around zero, indicating that we were equally likely to over- or under-estimate the strength of symptom propagation. This generally remained true when reporting bias was introduced, indicating that our estimates weren’t biased towards overestimating the strength of symptom propagation. Slight exceptions were seen for true values of *α* ≥ 0.6, where the median errors were positive in the case of secondary and primary-secondary reporting bias (blue squares and purple diamonds, respectively). However, the median errors were still relatively small (at most 0.09). Therefore, reporting bias led us to overestimate strong symptom propagation on average (although by a proportionately small amount).

In Supporting Information, we additionally show that, in the case of either secondary or primary-secondary reporting bias, the median error in *ν̂* was positive, indicating that *ν̂* was an overestimate of *ν* (Section S2.1, Fig. S1). This suggests that we are able to maintain our accurate *α* estimates because the error induced by the reporting bias is absorbed by our auxiliary parameter, *ν*. We found that the median errors in *ν* were generally close to zero under the no reporting bias and primary reporting bias scenarios.

We were also interested in whether there is evidence of symptom propagation relative to the null hypothesis that there is no symptom propagation (as typically assumed). The omission of symptom propagation mechanisms (were they truly present) can affect epidemiological modelling outcomes and, as a consequence, could impact public health intervention decisions [4]. With this motivation, we compared the effect of the different reporting bias scenarios on the percentage of replicates with support for symptom propagation (i.e. with *α* = 0 not in the 95% CI; Fig. 3, bottom row).

We found that, across data availability and reporting bias scenarios, the percentage of replicates in which symptom propagation was supported increased in an s-shaped fashion with *α*, i.e. as the true strength of symptom propagation increases, it becomes easier to detect. Further, the likelihood of detecting symptom propagation was effectively 100% for strengths above *α* = 0.2 once we had 1,000 pairs, irrespective of reporting bias (Fig. 3(f)). Even for 100 case pairs, *α* values above 0.3 were detected in over half of all replicates (Fig. 3(e)). In contrast, across all reporting bias scenarios, we found that for *n* = 10, there was no value of *α* for which there was support for symptom propagation across all replicates (Fig. 3(d)). In general, to be confident of not failing to detect symptom propagation, our results suggest that samples should aim for at least 100 case pairs, and ideally closer to 1,000.

Furthermore, false positives were rare. Across values of *n*, when *α* = 0, there was almost always no support for symptom propagation (i.e. *α* = 0 was consistently in the 95% CR). Even in the worst case of *n* = 10 and secondary or primary-secondary reporting bias, only 1.7% of replicates led to false positives.

#### 3.1.3 Accuracy of age-dependent estimates for the strength of symptom propagation

As explained in the Methods, we generated age-dependent synthetic data by assuming that age affected an individual’s symptom severity (by allowing *ν* to be age-dependent) and overall social mixing (meaning that individuals are more likely to interact with others of a similar age). We show in Supporting Information Section S3.1 that applying the age-free method (used in the previous section) to this age-dependent synthetic data led to overestimating the strength of symptom propagation and finding support for symptom propagation even when it did not exist (i.e. true *α* = 0). Specifically, for *n* = 1, 000 and *α* = 0, we observed a median error of 0.09 and erroneously found support for symptom propagation in 68% of replicates.

Here we show that extending the previously used methodology to estimate age-dependent *ν* values alongside a single value for the strength of symptom propagation effectively mitigates these effects and restores good estimates (Fig. 4). Our findings reflected those seen for the age-free results, with the range of our errors decreasing with *n*. For *n* = 1, 000, our estimates were consistently accurate (95% uncertainty interval of the error remained between −0.056 and 0.064).

**Fig. 4.**
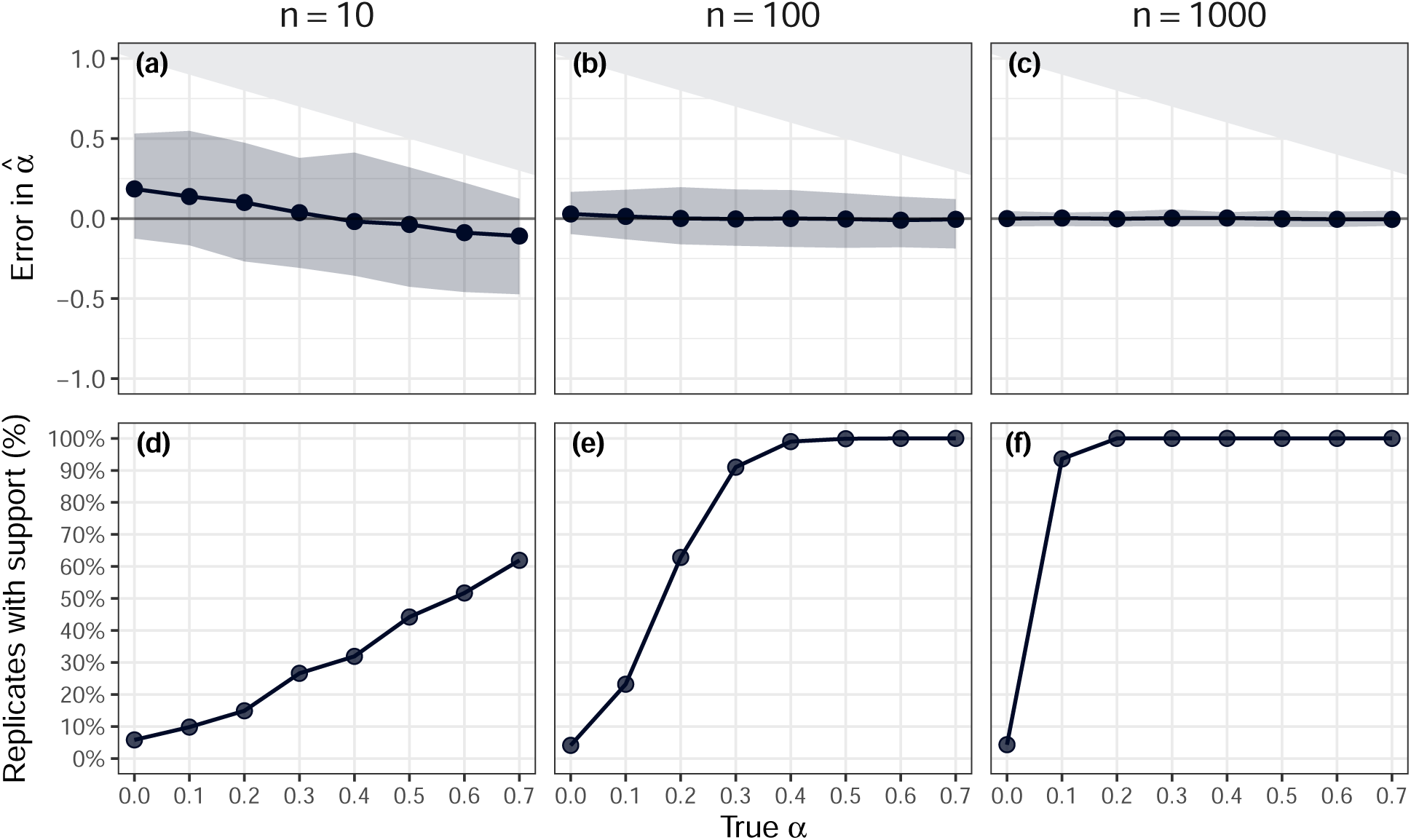
Error in the estimated values of *α̂* and the percentage of replicates with support for symptom propagation, using an age-dependent methodology. The top row shows the error in the estimates of *α*. A positive error corresponds to the estimated value being an overestimate of the true value. The solid grey regions correspond to infeasible errors, which are present due to the restriction of *α̂* being between 0 and 1. The points show the median errors and the lines show the interpolation between those points. The shaded area spans the 2.5th percentile to the 97.5th percentile (the 95% uncertainty interval). The bottom row gives the percentage of replicates (out of 1000) for which *α* = 0 was not within the 95% confidence region (i.e. support for symptom propagation). Across panels, we vary the assessed sample sizes *n* (i.e. the number of primary-secondary case pairs used in each replicate): **(a, d)** 10, **(b, e)** 100, **(c, f)** 1,000.

For *n* = 100 and *n* = 1, 000, our median errors were consistently close to zero and the 95% uncertainty intervals were symmetrical around an error of zero, indicating that we were equally likely to over- or under-estimate *α*. However, for *n* = 10, we found slight median errors for more extreme true values of *α*, where small true *α* was more likely to be overestimated and large true *α* was more likely to be underestimated. However, this is purely an artefact of our estimates now being posterior means, rather than maximum likelihood estimates. We verify in Supporting Information Section S3.3 that this effect is still seen even when the MCMC method is used to estimate age-free parameters from synthetic data with no age-structure.

In the Supporting Information, we additionally show the errors in the age-dependent *ν* values alongside the errors in *α* for comparison (Section S3.4, Fig. S5). We found that for large *n* (= 1, 000), the median errors across all three *ν̂* estimates were close to zero and the uncertainty intervals were narrow. However, for smaller *n*, the uncertainty intervals were wider and *ν*_1_ (the smallest of the three *ν* values) was overestimated whereas *ν*_3_ (the largest of the three *ν* values) was underestimated.

Using the age-dependent methodology, we found support for symptom propagation in only approximately 5% of replicates when the true value of *α* = 0 (5.8% for *n* = 10, 4.1% for *n* = 100, and 4.3% for *n* = 1000; Fig. 4, bottom row). This aligns with the fact that we are now considering 95% credible intervals, for which we would expect the true value to lie in that interval 95% of the time.

### 3.2 Real-world data results

We next produce estimates for the strength of symptom propagation from three real-world data sets from the COVID-19 pandemic. We first apply our age-free methodology to aggregate data on the number of primary-secondary case pairs with each possible combination of asymptomatic or symptomatic (a total of four combinations). Then we apply our age-dependent methodology to the same data sets, but now stratified by the age group of the secondary case (noting that the Norway contact tracing data was omitted from this analysis due to not having age group data available).

#### 3.2.1 Age-free results

We applied the age-free method to obtain maximum likelihood estimates for the strength of symptom propagation, *α*, and the baseline probability of severe disease, *ν*. The estimates and 95% confidence regions are shown in Fig. 5 with values given in Table 4.

**Fig. 5.**
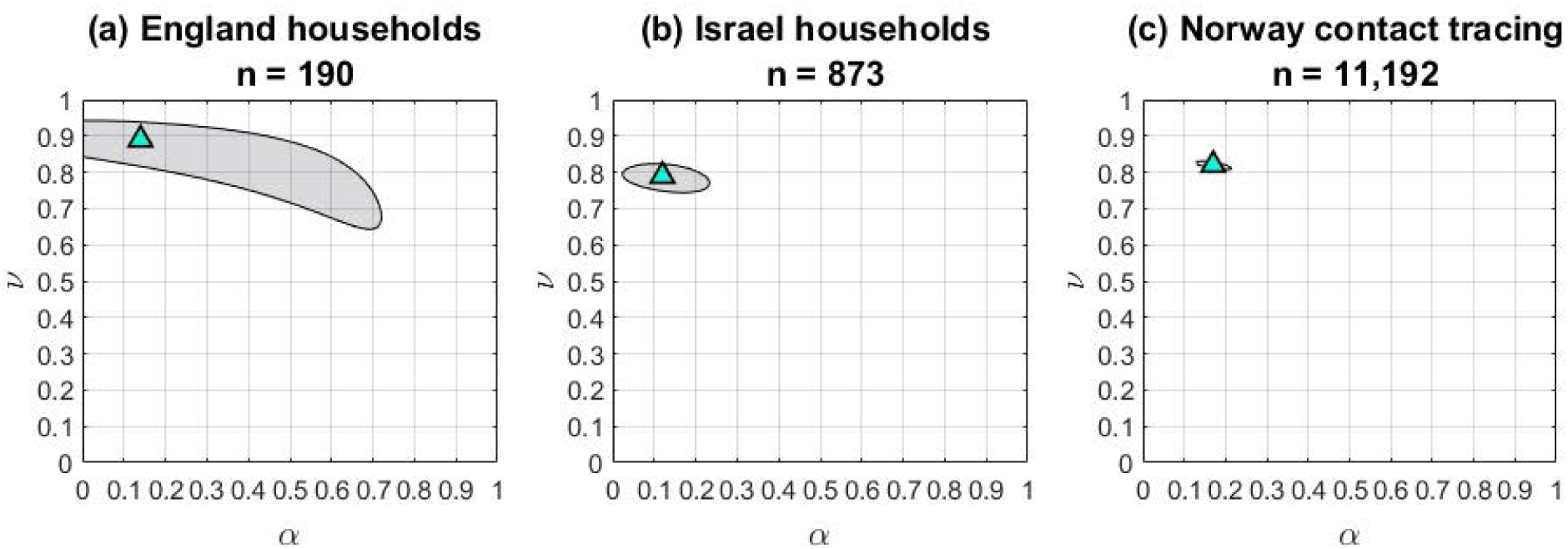
95% bivariate confidence regions for three real-world data sets. 95% confidence regions for the predicted value of *ν* and *α* for the three real-world data sets: **(a)** England households, **(b)** Israel households, **(c)** Norway contact tracing. Here *n* is the number of primary-secondary case pairs in the data set. The cyan triangles indicate the MLE estimates for each replicate, and the grey shaded regions show the 95% bivariate confidence regions.

**Table 4.**
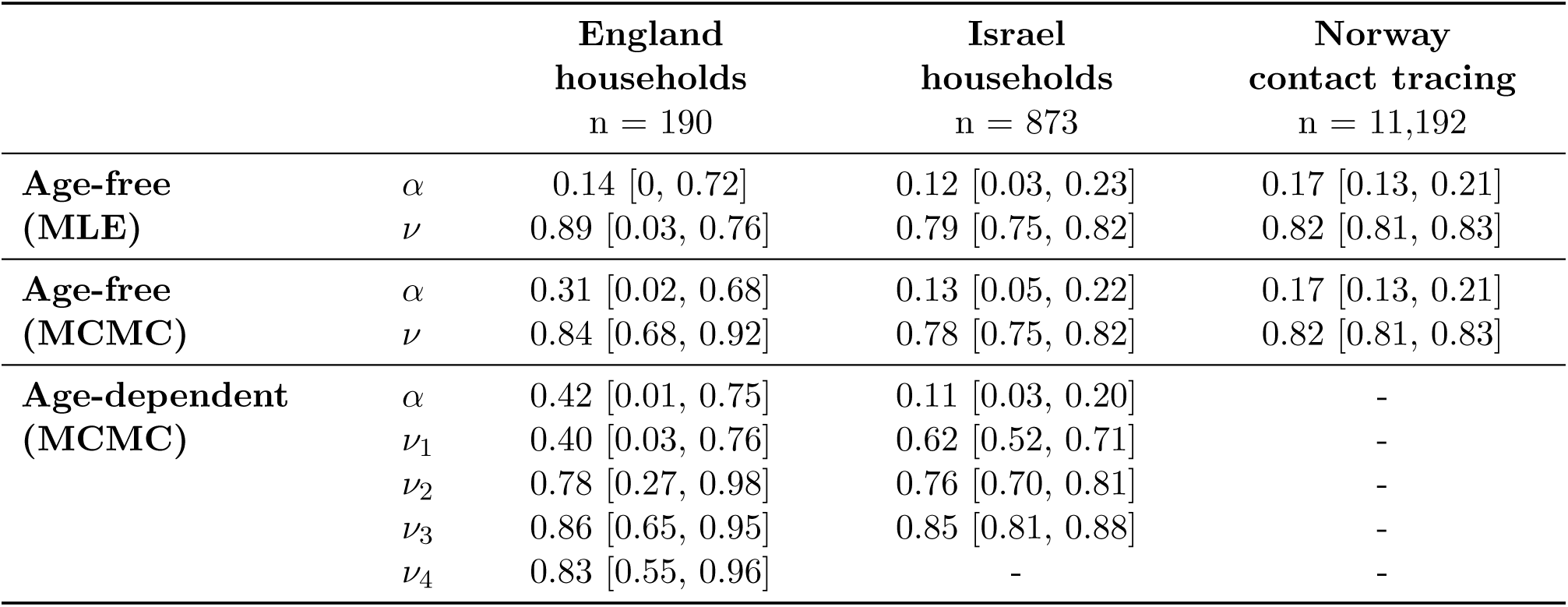
Estimates for. *α* **and** *ν* **for two real-world data sets, comparing age-free vs age-dependent results.** Here we give the values of the MLEs and their 95% confidence intervals for the age-free (MLE) estimates and the posterior means and 95% credible intervals for the age-free (MCMC) and age-dependent (MCMC) estimates. Note that the age groups corresponding to each age-dependent *ν* estimate vary between the two household data sets: 0–10, 11–18, 19–54 and 55+ for England, and 0–9, 10–19 and 20+ for Israel. Here *n* is the number of primary-secondary case pairs in the data set.

|  |  | England<br>households<br>n = 190 | Israel<br>households<br>n = 873 | Norway<br>contact tracing<br>n = 11,192 |
| --- | --- | --- | --- | --- |
| **Age-free**<br>**(MLE)** | $\alpha$ | 0.14 [0, 0.72] | 0.12 [0.03, 0.23] | 0.17 [0.13, 0.21] |
| | $\nu$ | 0.89 [0.03, 0.76] | 0.79 [0.75, 0.82] | 0.82 [0.81, 0.83] |
| **Age-free**<br>**(MCMC)** | $\alpha$ | 0.31 [0.02, 0.68] | 0.13 [0.05, 0.22] | 0.17 [0.13, 0.21] |
| | $\nu$ | 0.84 [0.68, 0.92] | 0.78 [0.75, 0.82] | 0.82 [0.81, 0.83] |
| **Age-dependent**<br>**(MCMC)** | $\alpha$ | 0.42 [0.01, 0.75] | 0.11 [0.03, 0.20] | - |
| | $\nu_1$ | 0.40 [0.03, 0.76] | 0.62 [0.52, 0.71] | - |
| | $\nu_2$ | 0.78 [0.27, 0.98] | 0.76 [0.70, 0.81] | - |
| | $\nu_3$ | 0.86 [0.65, 0.95] | 0.85 [0.81, 0.88] | - |
| | $\nu_4$ | 0.83 [0.55, 0.96] | - | - |

Overall, parameter estimates were relatively consistent across the three data sets, with *α* being estimated within the range 0.12-0.17. Importantly, for both households in the Israel households and Norway contact tracing data sets (Fig. 5(b) and 5(c)), *α* = 0 was not in the 95% confidence region, indicating that these data sets provide support for the occurrence of symptom propagation for SARS-CoV-2. The estimates for *ν* were consistently high (at least 0.79), reflecting the low prevalence of asymptomatic cases in the data sets.

The size of the 95% confidence regions varied across the data sets, which is unsurprising given the differences in data availability. Consistently, there was greater uncertainty in *α* compared to *ν*. This was seen most obviously for the households in England (*n* = 190, Fig. 5(a)) where the uncertainty in *α* was notably greater than seen in our synthetic data analysis with *n* = 100 and true *α* = 0.2 (a width of 0.72 vs an average width of 0.43, Fig. 2(b)). This exacerbated uncertainty likely stems from the very small number of asymptomatic primary cases in the England households data, whereas in our synthetic data analysis we assumed half of all cases were mild. For the Israel households (*n* = 873, Fig. 5(b)) the uncertainty in *α* was higher than the average seen for *n* = 1, 000 and true *α* = 0.2 in the synthetic data analysis, but much less than that seen for *n* = 100 (0.26 vs 0.15 vs 0.43).

#### 3.2.2 Age-dependent results

We then applied the age-dependent methodology to two of the real-world data sets: households in England and in Israel, for which we also had access to age information about the primary and secondary cases. This methodology produced posterior distributions for *α* and age-dependent values of *ν* (*ν*_1_*, ν*_2_*, ν*_3_*, ν*_4_ for households in England, corresponding to the age classes 0–10, 11–18, 19–54 and 55+ and *ν*_1_*, ν*_2_*, ν*_3_ for households in Israel, corresponding to the age classes 0–9, 10–19 and 20+). The posterior distributions are shown in Fig. 6 and the posterior means and 95% credible intervals are listed in Table 4.

**Fig. 6.**
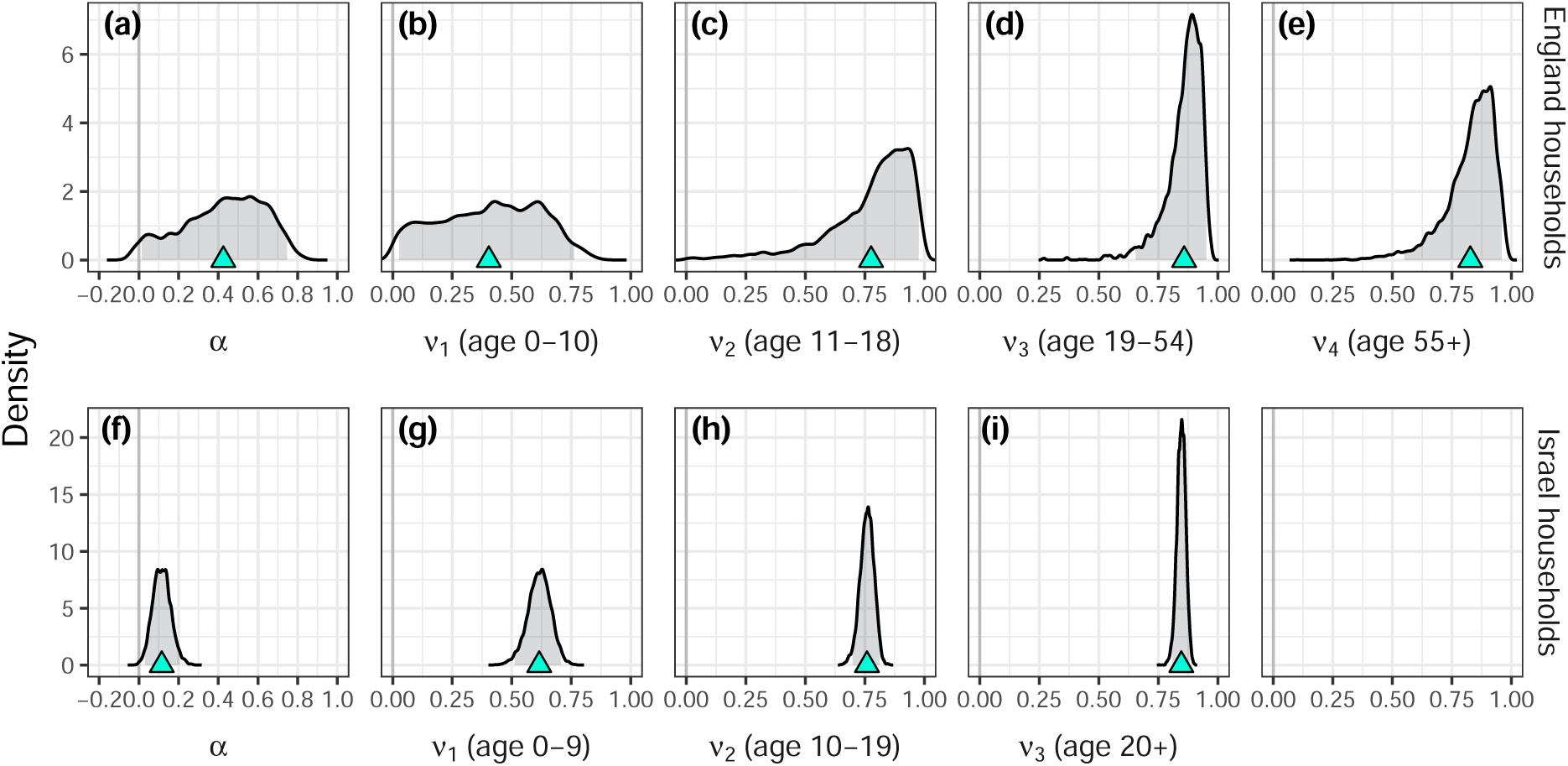
Posterior distributions for *α* and four age-dependent *ν* values, for two real-world data sets. Posterior distributions for *α* and the four age-dependent *ν* parameters for two real-world data sets: England households (top row) and Israel households (bottom row). The cyan triangles indicate the posterior means and the grey shaded regions indicate the 95% credible intervals. Note that the age groups corresponding to each age-dependent *ν* estimate vary between the two household data sets: 0-10, 11-18, 19-54 and 55+ for England, and 0-9, 10-19 and 20+ for Israel. Therefore, there is no *ν*_4_ parameter for the Israel households data.

For the Israel households data, our estimate for *α* was similar to that seen when the age-free method-ology was used (0.11 vs 0.12, Fig. 6(f)), indicating that our positive estimate for the strength of symptom propagation was not a result of age-dependent effects. In this case, we saw a single narrow peak in the posterior distribution that aligned with the posterior mean, highlighting the certainty of our estimate. For the England households data, we surprisingly found that our estimate for *α* increased when the age-dependent methodology was used (0.42 vs 0.14, Fig. 6(a)). We found that the posterior distribution and 95% credible interval was much wider than for Israel, which was expected due to the smaller number of pairs in the England data set. This high uncertainty may be the reason for the observed increase in our estimate of *α*.

Additionally, the small size of the England households data set, particularly when stratified by secondary case age group, likely caused the MCMC methodology to be less effective. We found that the chain was less well-mixed for the England data, with a lower acceptance rate of 0.174 (compared to 0.284 for the Israel data). The trace plots for this analysis are provided in Supporting Information Section S5.1 (Figs. S20 and S21).

As with the age-free results, our estimates for *ν* were consistently high. In particular, with the exception of the youngest age groups for each data set, *ν* was found to be at least 0.76. This is again likely to be due to reporting bias in the data. We also found that generally the value of *ν* increased with the secondary case age group (with the exception of *ν*_4_ (ages 55+) being slightly lower than *ν*_3_ (ages 19-54) for the England households data set, although the difference was minimal).

To demonstrate that our findings are not purely a result of using MCMC rather than MLE, we additionally estimated parameters using an age-free MCMC procedure. The posterior means and 95% credible intervals are listed in Table 4 alongside the age-free MLE estimates and age-dependent MCMC estimates. The posterior distributions are shown in the Supporting Information (Fig. S22)

We found that whether age-free MLE or age-free MCMC was used generally had a very minimal impact on our estimates, with the exception of England households (Table 4). In the case of the Norway contact tracing data, the central estimates were the same regardless of whether the age-free MLE or age-free MCMC method was used. Similarly, for Israel households, the variations seen were minor with the central estimates for both *α* and *ν* only varying by 0.01. In contrast, the estimates for England households varied quite substantially, with the age-free MCMC method producing a much higher estimate for *α* than MLE (0.31 vs 0.14). This increase in our estimate is likely due to *α̂* now being the mean of the posterior distribution, which was relatively flat across values of *α* due to the minimal data availability (Fig. S22(a)). In particular, the posterior density was high for all values of *α* between 0.1 and 0.5. The switch from MLE to MCMC may partially explain why we found a higher estimate for *α* when the age-dependent methodology was used.

## 4 Discussion

In this paper, we introduced a method to estimate the strength of symptom propagation from summary data on primary-secondary case pairs and their symptom presentations. We used synthetic data to explore the effect of differing data availability on our estimates of these parameters and verify the accuracy of our methodology. We additionally considered the effect of reporting bias. The method can be extended to account for other factors that affect severity, which we demonstrated using age as an example. Then, we applied our methodology to generate estimates for the strength of symptom propagation for SARS-CoV-2 for three example real-world datasets from the COVID-19 pandemic: household data from England [27] and Israel [28], and contact tracing data from Norway [6].

Our analysis of these datasets using our age-free methodology returned consistent estimates of the strength of symptom propagation for SARS-CoV-2 (*α* = 0.14, 0.12, 0.17), meaning that a secondary case would be 12-17% more likely to be symptomatic if the primary case was symptomatic versus the primary case being asymptomatic. Putting this finding into context, a meta-analysis for SARS-CoV-2 estimated the proportion of infections that were symptomatic across age groups to be approximately 53% for 0-18, 68% for 19-59, and 80% for 60+ [33]. Therefore, the increase in risk of symptomatic infection resulting from symptom propagation is of a similar magnitude to the increase in risk by moving up an age group (a 15% increase in risk comparing 19-59 to 0-18 and a 12% increase in risk comparing 60+ to 19-59). We also applied our age-dependent methodology to the England and Israel households data sets (for which age group data were available). Our positive estimates for the strength of symptom propagation persisted (*α* = 0.42, 0.11). For both datasets, *α* = 0 was not in the 95% confidence interval, indicating significant support for symptom propagation. These findings suggest that the positive estimates of symptom propagation obtained without accounting for age structure are not driven by age-dependent effects.

Our estimates of symptom propagation aligned with previous studies. Santermans *et al.* [5] estimated symptom propagation-like parameters from 2009 H1N1 incidence data using the difference in contact patterns between asymptomatic and symptomatic cases. They estimated that the probability of being symptomatic if infected by a symptomatic index case was 0.22 vs 0.02 if infected by an asymptomatic index (this is an increase in risk of 0.2, so is equivalent to *α* = 0.2). Also giving support for symptom propagation was the study of COVID-19 by Methi and Madslien [6] from which we sourced the Norway contact tracing data. Whilst the authors did not specifically estimate the strength of symptom propagation, they found that the odds of a secondary case being asymptomatic were significantly higher if the primary case was asymptomatic (OR 2.67, 95% CI [2.28, 3.12]). Furthermore, correlations in symptom presentation persisted after accounting for confounding factors, which included the age of both the primary and secondary cases.

We validated our methodology by using synthetic data to assess accuracy and data requirements. We found that we only required a relatively small number of primary-secondary case pairs to obtain accurate estimates for the strength of symptom propagation. Specifically, 1,000 pairs was sufficient for errors to be consistently small across 1,000 replicates (at most underestimating *α* by 0.06 and at most overestimating *α* by 0.07). Our methodology retained high accuracy even when severity-dependent reporting biases were present in the data, which are commonly observed in household and contact tracing studies due to self-selecting volunteers and/or participants self-reporting their contacts [8]. Verifying our methodology retained high accuracy in the presence of severity-dependent reporting biases was particularly important due to the small number of asymptomatic cases in our three real-world datasets. Finally, we verified that we very rarely found false positives (support for symptom propagation when it was not present in the underlying data). False positives occurred in at most 1.3% of synthetic replicates across data availability and reporting bias assumptions.

Our approach provides advantages over those used in the existing literature. For example, a limitation of Santermans *et al.* [5] was that they could not estimate both the symptom propagation parameters and the relative transmissibility of symptomatic and asymptomatic cases. This unidentifiability is not an issue when using contact tracing data and our method – our likelihood functions involve only *α* and *ν*, with no additional assumptions required for the other parameters. Furthermore, we demonstrated that reliable estimates can be produced using straightforward methodologies and extremely modest data requirements. For the age-free method, we require only four summary statistics on the number of primary-secondary case pairs of each combination of symptom presentations. The extension of our methodology to include age-structure only requires four summary statistics per secondary case age group, corresponding to the number of primary-secondary case pairs of each combination of symptom presentation for each secondary case age group, with no data needed on the ages of primary cases or on social mixing patterns. This is a particularly important benefit since raw contact tracing data often cannot be shared due to individuals being identifiable [34, 35].

A limitation of our real-world data analysis was that two of the data sets consisted of purely household data, as opposed to contact tracing data. Household data has higher uncertainty regarding who infected whom, or even whether the secondary case resulted from within-household transmission. However, we would expect that this would bias our results to lower estimates for the strength of symptom propagation, as individuals who are not in the same chain of transmission should not have correlated symptoms. This problem could be potentially mitigated by using genome sequencing data, which has previously been used to determine who infected whom for other pathogens [36, 37]. An additional issue likely to arise from household data is that many individuals are likely to be genetically related. These genetic similarities may cause correlations in symptom severity [38, 39]. Relevant mitigations include restricting analysis to two-person households or settings where individuals are less likely to be related (e.g. a university [40]).

Another limitation is that other characteristics beyond age could cause biases in our estimates for the strength of symptom propagation. Variants of COVID-19 are intrinsically correlated between primary and secondary cases and often affect symptom severity [41]. Furthermore, vaccination often acts to reduce disease severity [42] and a range of socioeconomic factors mean that vaccinated individuals are likely to be socially clustered [43]. However, these factors could not impact our strength of symptom propagation estimates for SARS-CoV-2 obtained from the Israel households dataset. Those data were collected at the very beginning of the pandemic (between 17^th^ March and 3^rd^ May 2020) when only the wild-type SARS-CoV-2 variant was in circulation and there was not yet a vaccine for SARS-CoV-2. In contrast, the England households dataset was collected between February and August 2021, when both the Alpha and Delta variants were in circulation and a vaccination campaign was underway. The presence of these factors could contribute to our estimate for the strength of symptom propagation being higher for England than for Israel households. In the same way that age groups were accounted for, our methodology could straightforwardly be extended to account for both variant and vaccination status if this information was available. Given sufficient data availability, the methodology can plausibly account for all three factors at once by extending *ν* to be a three dimensional matrix of values.

In summary, we have demonstrated that the strength of symptom propagation can be robustly estimated using data on primary-secondary case pairs, even with data on relatively few cases. Our methodology is robust to reporting bias. It can be extended to incorporate key determinants of disease severity, such as age structure, without requiring data on primary case characteristics or social mixing patterns. Applying this methodology to real-world data sets from the COVID-19 pandemic provided consistent evidence that symptom propagation occurs for SARS-CoV-2, underscoring its epidemiological relevance. Our approach provides a practical tool for investigating symptom propagation across a wide range of pathogens and settings. Crucially, given its minimal data requirements and flexibility, it is well-suited for enabling the estimation of the strength of symptom propagation in situations where large-scale data sets are unavailable or cannot be shared.

## Author contributions

**Phoebe Asplin:** Conceptualisation, Data curation, Formal analysis, Investigation, Methodology, Software, Validation, Visualisation, Writing - Original Draft, Writing - Review & Editing.

**Rebecca Mancy:** Conceptualisation, Investigation, Methodology, Supervision, Visualisation, Writing - Review & Editing.

**Matt J. Keeling:** Conceptualisation, Investigation, Methodology, Supervision, Visualisation, Writing - Review & Editing.

**Edward M. Hill:** Conceptualisation, Investigation, Methodology, Software, Supervision, Validation, Visualisation, Writing - Original Draft, Writing - Review & Editing.

## Data availability

All data utilised in this study are publicly available, with relevant references and data repositories provided.

## Code availability

The code repository for the study is available at:

https://github.com/pasplin/symptom_propagation_estimation.git. Archived code: https://doi.org/10.5281/zenodo.19368583.

