## Supporting Information for "Estimating the strength of symptom propagation from primary-secondary case pair data"

### Supporting Information
### Estimating the strength of symptom propagation from data on primary-secondary case pairs

Phoebe Asplin[1,2*], Rebecca Mancy[3,4], Matt J. Keeling[2,5], Edward M. Hill[6,7,8,9*]

**1** EPSRC & MRC Centre for Doctoral Training in Mathematics for Real-World Systems, University of Warwick, Coventry, United Kingdom.
**2** The Zeeman Institute for Systems Biology & Infectious Disease Epidemiology Research, University of Warwick, Coventry, United Kingdom.
**3** School of Biodiversity, One Health and Veterinary Medicine, University of Glasgow, Glasgow, United Kingdom.
**4** Wales Applied Virology Unit (WAVU), School of Medicine, Cardiff University, Cardiff, United Kingdom.
**5** Mathematics Institute and School of Life Sciences, University of Warwick, Coventry, United Kingdom.
**6** Civic Health Innovation Labs, University of Liverpool, Liverpool, United Kingdom.
**7** Department of Public Health, Policy and Systems, Institute of Population Health, University of Liverpool, Liverpool, United Kingdom.
**8** NIHR Health Protection Research Unit in Emerging and Zoonotic Infections, University of Liverpool, Liverpool, United Kingdom.
**9** The Pandemic Institute, Liverpool, United Kingdom.

#### S1 Additional methods

Here we provide some additional details of our synthetic data generation and parameter estimation. First we describe how we fixed the overall proportion of cases that were severe when generating age-free synthetic data (Section S1.1). Then we provide the algorithms used for generating both the age-free and age-dependent synthetic data sets (Sections S1.2 and S1.3). We then describe how the age-free and age-dependent likelihoods were derived (Section S1.4). Finally, we provide additional details of how MCMC was applied to generate our age-dependent estimates for the symptom propagation parameters (Section S1.5).

##### S1.1 Fixing the proportion of cases that are severe

We have the following equations for the SIR ODE model with symptom propagation.

$$\begin{aligned}
\frac{dS}{dt} &= -(\lambda_m + \lambda_s)S, \\
\frac{dI_m}{dt} &= \Big(\big(\alpha + (1-\alpha)(1-\nu)\big)\lambda_m + (1-\alpha)(1-\nu)\lambda_s\Big)S - \gamma_m I_m, \\
\frac{dI_s}{dt} &= \Big((1-\alpha)\nu\lambda_m + \big(\alpha + (1-\alpha)\nu\big)\lambda_s\Big)S - \gamma_s I_s, \\
\frac{dR}{dt} &= \gamma_m I_m + \gamma_s I_s),
\end{aligned} \tag{1}$$

where the force of infection from mild infections, $\lambda_m$, and severe infections, $\lambda_s$, respectively, are given by:

$$\lambda_m = \frac{\beta_m I_m}{N}, \quad \lambda_s = \frac{\beta_s I_s}{N},$$

where $N$ is the population size that I assumed to be constant.

Let $\rho(t) = \frac{I_s(t)}{I_m(t)}$ be the ratio of severe to mild infections at time $t$. Using the quotient rule, I derived a differential equation for the rate of change of $\rho$,

$$\begin{aligned}
\frac{d\rho}{dt} =& \frac{I_m(t)\frac{dI_s}{dt} - I_s(t)\frac{dI_m}{dt}}{I_m(t)^2} \\
=& \frac{1}{I_m(t)^2 N}\Bigg[I_m(t)\Big((1-\alpha)\nu\beta_m S(t)I_m(t) + \big(\alpha+(1-\alpha)\nu\big)\beta_s S(t)I_s(t) - \gamma_s I_s(t)\Big) \\
& - I_s(t)\Big(\big(\alpha+(1-\alpha)(1-\nu)\big)\beta_m S(t)I_m(t) + (1-\alpha)(1-\nu)\beta_s S(t)I_s(t) - \gamma_m I_m(t)\Big)\Bigg].
\end{aligned}$$

Substituting in $\rho(t) = \frac{I_s(t)}{I_m(t)}$,

$$\begin{aligned}
\frac{d\rho}{dt} =& \Big((1-\alpha)\nu\beta_m + \big((\alpha+(1-\alpha)\nu)\beta_s - (\alpha+(1-\alpha)(1-\nu))\beta_m\big)\rho(t) - (1-\alpha)(1-\nu)\beta_s\rho(t)^2\Big)\frac{S(t)}{N} \\
& + (\gamma_m - \gamma_s)\frac{\rho(t)}{N}.
\end{aligned}$$

Note that in the case where $\gamma_m \neq \gamma_s$, the value of $\rho(t)$ such that $\frac{d\rho}{dt} = 0$ depends on $S(t)$. Therefore, we would need to know how $S(t)$ varies over time to calculate an equilibrium value of $\rho(t)$.

However, if $\gamma_m = \gamma_s$,

$$\frac{d\rho}{dt} = \Big((1-\alpha)\nu\beta_m + \big((\alpha + (1-\alpha)\nu)\beta_s - (\alpha + (1-\alpha)(1-\nu))\beta_m\big)\rho(t) - (1-\alpha)(1-\nu)\beta_s\rho(t)^2\Big)\frac{S(t)}{N}. \tag{2}$$

Therefore, $\rho$ is at an equilibrium, $\rho^*$, when

$$(1-\alpha)\nu\beta_m + \big((\alpha + (1-\alpha)\nu)\beta_s - (\alpha + (1-\alpha)(1-\nu))\beta_m\big)\rho^* - (1-\alpha)(1-\nu)\beta_s\rho^{*2} = 0. \tag{3}$$

Assuming that $\alpha \in [0, 1)$ and $\nu \in [0, 1)$,

$$\begin{aligned}\rho^*_\pm =& \frac{1}{2(1-\alpha)(1-\nu)\beta_s}\Bigg[\big((\alpha + (1-\alpha)\nu)\beta_s - (\alpha + (1-\alpha)(1-\nu))\beta_m\big) \\ &\pm \sqrt{\big((\alpha + (1-\alpha)\nu)\beta_s - (\alpha + (1-\alpha)(1-\nu))\beta_m\big)^2 + 4\nu(1-\alpha)^2(1-\nu)\beta_m\beta_s}\Bigg].\end{aligned}$$

Notice that the square root term is real, since

$$\begin{aligned}&\big((\alpha + (1-\alpha)\nu)\beta_s - (\alpha + (1-\alpha)(1-\nu))\beta_m\big)^2 + 4\nu(1-\alpha)^2(1-\nu)\beta_m\beta_s \\ &\geq \big((\alpha + (1-\alpha)\nu)\beta_s - (\alpha + (1-\alpha)(1-\nu))\beta_m\big)^2 \\ &\geq 0.\end{aligned}$$

Furthermore, note that $\rho^*_+$ is non-negative (and $\rho^*_-$ is non-positive) because

$$\begin{aligned}&\sqrt{\big((\alpha + (1-\alpha)\nu)\beta_s - (\alpha + (1-\alpha)(1-\nu))\beta_m\big)^2 + 4\nu(1-\alpha)^2(1-\nu)\beta_m\beta_s} \\ &\geq \big((\alpha + (1-\alpha)\nu)\beta_s - (\alpha + (1-\alpha)(1-\nu))\beta_m\big).\end{aligned}$$

Therefore, there always exists a single real, non-negative equilibrium,

$$\begin{aligned}\rho^* =& \frac{1}{2(1-\alpha)(1-\nu)\beta_s}\Bigg[\Big((\alpha + (1-\alpha)\nu)\beta_s - (\alpha + (1-\alpha)(1-\nu))\beta_m\Big) \\ &+ \sqrt{\big((\alpha + (1-\alpha)\nu)\beta_s - (\alpha + (1-\alpha)(1-\nu))\beta_m\big)^2 + 4\nu(1-\alpha)^2(1-\nu)\beta_m\beta_s}\Bigg].\end{aligned} \tag{4}$$

Consequently, there exists a ratio of severe to mild infections for which if the outbreak starts in this ratio, then the ratio (and therefore the proportion of infections that are severe) will remain constant throughout the whole outbreak.

Writing $\beta_m = \beta$ and $\beta_s = b\beta$,

$$\begin{aligned}\rho^* =& \frac{1}{2b(1-\alpha)(1-\nu)}\Bigg[\Big((\alpha(1-\nu) + \nu)b - (\alpha\nu + (1-\nu))\Big) \\ &+ \sqrt{\big((\alpha(1-\nu) + \nu)b - (\alpha\nu + (1-\nu))\big)^2 + 4\nu(1-\alpha)^2(1-\nu)b}\Bigg].\end{aligned} \tag{5}$$

So $\rho^*$ only depends on $\alpha, \nu$ and $b$. Note that, in particular, $\rho^*$ does not depend on $\beta$.

Rearranging this equation gives an equation for $\nu$,

$$\nu = \frac{\rho(1 + b - 2\alpha b)}{(1-\alpha)(1+\rho)(1+b\rho)},$$

Across all values of $\alpha$, we chose a value of $\nu$ to fix the proportion of cases that are severe, P to be = 0.5. Equivalently, we assumed equal numbers of severe and mild cases, i.e. the ratio of severe to mild cases is $\rho = 1$. In this study, we assume that the overall proportion of cases that are severe is equal to the equilibrium value (i.e. P = P$^*$ and consequently $\rho = \rho^*$).

Substituting $b = \beta_s/\beta_m = 2$ and $\rho = 1$, we obtain

$$\nu = \frac{3 - 4\alpha}{6(1-\alpha)}.$$

Note that as $\alpha$ increases, the value of $\nu$ required to give the fixed value of P decreases and at some point becomes negative. It can be calculated that, for our parameterisation, the maximum possible value of $\alpha$ that allows for non-negative $\nu$ is 0.75. As we studied values of $\alpha$ in increments of 0.1, we therefore considered a maximum value of $\alpha = 0.7$.

#### S1.2 Algorithm for generating age-free synthetic data

We generated the synthetic data using the $\tau$-leap approximation for the Gillespie algorithm [44] (Algorithm 1) with a timestep of $\tau = 1$ day. This approximation was used as it is much faster than the exact method, which is important due to us assuming a population size of $N = 100,000$ individuals, akin to a small city. This population size was chosen to balance two goals: ensuring there were enough cases so that samples for each of the two data streams could be considered independent, whilst keeping the population size small enough that assuming everyone could potentially interact remained realistic (as is assumed in the Gillespie algorithm). As this timestep is relatively large, it is possible that this approximation could cause errors in the simulated epidemic. However, these errors would have a most notable effect when case numbers are small (i.e. at the start and end of the epidemic) and these cases are unlikely to be included in the sampled data set, since cases from each timestep are sampled in proportion to the number of cases occurring at that time.

From the synthetic data generation process, we output primary-secondary case pair data, akin to contact tracing data. The contact tracing data outputs are $C^{m\to m}, C^{m\to s}, C^{s\to m}, C^{s\to s}$, the total numbers of each type of infection event (i.e. counts of events of mild case generating a mild case, mild case generating a severe case, severe case generating a mild case, severe case generating a severe case).

We initialised the simulation with one mild infected individual and one severe infected individual ($I_m(0) = I_s(0) = 1$). We assumed there was no prior immunity in the population (such that there were not recovered individuals; $R(0) = 0$), so all other individuals were susceptible ($S(0) = 100,000 - 2$). We generated a single synthetic dataset under these conditions for each considered value of $\alpha$, ranging from 0 to 0.7 in increments of 0.1.

**Algorithm 1** Generating synthetic case data for a population of 99,998 individuals with initial conditions $\{S(0), I_m(0), I_s(0), R(0)\} = \{100,000-2, 1, 1, 0\}$. we used a time step of $\tau = 1$ day. We output aggregate contact tracing data, $C^{m\to m}, C^{m\to s}, C^{s\to m}, C^{s\to s}$ (the number of infection events with each possible combination of primary-secondary case severity).

```
Input: T_end, {S(0), I_m(0), I_s(0), R(0)}, τ
{S, I_m, I_s, R} ← {S(0), I_m(0), I_s(0), R(0)}        ▷ Set up compartment initial conditions
C^{m→m}, C^{m→s}, C^{s→m}, C^{s→s} ← 0                  ▷ Initialise output vectors
t ← 0
while t < T_end do
    K^{m,m} ← Poisson(τ β_m S I_m (α + (1 − α)(1 − ν)))  ▷ Number of infection events m → m
    K^{m,s} ← Poisson(τ β_m S I_m (1 − α)ν)              ▷ Number of infection events m → s
    K^{s,m} ← Poisson(τ β_s S I_s (1 − α)(1 − ν))        ▷ Number of infection events s → m
    K^{s,s} ← Poisson(τ β_s S I_s (α + (1 − α)ν))        ▷ Number of infection events s → s
    K^m ← Poisson(τ γ_m I_m)                             ▷ Number of mild recovery events
    K^s ← Poisson(τ γ_s I_s)                             ▷ Number of severe recovery events
    for X ∈ {m, s} do
        for Y ∈ {m, s} do
            C^{X→Y} ← C^{X→Y} + min(P^{X,Y}, S)
            if Y = m then
                I_m ← I_m + min(K^{X,Y}, S)
            else if Y = s then
                I_s ← I_s + min(K^{X,Y}, S)
            end if
            S ← max(S − K^{X,Y}, 0)
        end for
    end for
    I_m ← max(I_m − K^m, 0)
    R ← R + min(K^m, I_m)
    I_s ← max(I_s − P^s, 0)
    R ← R + min(K^s, I_s)
    t ← t + τ
end while
Return: C^{m→m}, C^{m→s}, C^{s→m}, C^{s→s}
```

#### S1.3 Algorithm for generating age-structured synthetic data

To run stochastic simulations of the age-structured symptom propagation model, we again used the $\tau$-leap approximation for the Gillespie algorithm.

Working with an overall population size of $N = 100,000$ individuals, we assumed 20,000 were aged 0-17, 60,000 were aged 18-64 and 20,000 were 65+. We assumed the outbreak started with one mild and one severe case in each age group, and all other individuals were susceptible. We generated a single synthetic dataset under these conditions for each considered value of $\alpha$, ranging from 0 to 0.9 in increments of 0.1. Note that as we are no longer fixing the proportion of cases that are severe, there is no requirement to have $\alpha \leq 0.75$.

In order to simulate realistic social mixing, we used a social contact matrix $M$ that determined the rate at which individuals mix with those from the same age class versus the other age classes. The rates were based on the POLYMOD social mixing survey, using data for the UK from 2008 [14]. We used the R package *socialmixr* to aggregate the values into our desired age groups. We rescaled the resulting contact matrix to give a maximum eigenvalue of one so that the overall rate at which people mix was kept constant.

$$M = \begin{bmatrix} 0.66 & 0.17 & 0.06 \\ 0.49 & 0.69 & 0.34 \\ 0.05 & 0.09 & 0.15 \end{bmatrix}$$

.

This synthetic data generation procedure output age-stratified aggregate contact tracing data:

$$C_i^{m \to m}, C_i^{m \to s}, C_i^{s \to m}, C_i^{s \to s},$$

the total numbers of each type of infection event where the secondary case was in age group $i$ (i.e. counts per time step of events of mild case generating a mild case, mild case generating a severe case, severe case generating a mild case, severe case generating a severe case).

#### S1.4 Derivation of the likelihood of the symptom propagation parameters

We derived likelihoods from the sampled primary-secondary case pair data to allow us to obtain parameter estimates for $\alpha$ and $\nu$. We first consider the age-free scenario, where we estimate $\alpha$ and a single value of $\nu$ from age-free data using maximum likelihood estimation. We then consider the age-dependent scenario where we estimate $\alpha$ and three age-dependent values of $\nu$ from age-structured data using Markov Chain Monte-Carlo (MCMC, rather than maximum likelihood estimation) since we are now estimating five parameters (compared to only two for the age-free methodology) which would make maximum likelihood estimation very computationally expensive.

##### Age-free likelihood

We have a sample of aggregate contact tracing data, which describes the number of primary-secondary case pairs with each possible combination of symptom severities: $\tilde{C}^{m \to m}, \tilde{C}^{m \to s}, \tilde{C}^{s \to m}, \tilde{C}^{s \to s}$.

The model framework described in Section 2.1 specifies the probability that, given the primary case has severity $X$, the secondary case has severity $Y$, which we denote $\mathbb{P}(X \to Y)$ (see Fig. 1). Given

**Algorithm 2** Generating synthetic case data for a population of 100,000 individuals with initial conditions $\{S_i(0), I_{m,i}(0), I_{s,i}(0), R_i(0)\} = \{N_i - 2, 1, 1, 0\}$. We used a time step of $\tau = 1$ day. We output aggregate contact tracing data, $C_i^{m\to m}, C_i^{m\to s}, C_i^{s\to m}, C_i^{s\to s}$ (the number of infection events with each possible combination of primary-secondary case severity) where $i$ is the age group of the secondary case.

---

**Input:** $T_{\text{end}}, \{\mathbf{S}(0), \mathbf{I}_m(0), \mathbf{I}_s(0), \mathbf{R}(0)\}, \tau$
$\{\mathbf{S}, \mathbf{I}_m, \mathbf{I}_s, \mathbf{R}\} \leftarrow \{\mathbf{S}(0), \mathbf{I}_m(0), \mathbf{I}_s(0), \mathbf{R}(0)\}$ ▷ Set up compartment initial conditions
$\mathbf{C}^{m\to m}, \mathbf{C}^{m\to s}, \mathbf{C}^{s\to m}, \mathbf{C}^{s\to s} \leftarrow \mathbf{0}$ ▷ Initialise output vectors
$t \leftarrow 0$
**while** $t < T_{\text{end}}$ **do**
  **for** age group $i$ **do**
    $K_i^{m,m} \leftarrow \text{Poisson}\Big(\tau \sum_{j=1}^{A} M(i,j)\beta_m I_{m,j} S_i(\alpha + (1-\alpha)(1-\nu_i))\Big)$ ▷ Infection $m \to m$
    $K_i^{m,s} \leftarrow \text{Poisson}\Big(\tau \sum_{j=1}^{A} M(i,j)\beta_m I_{m,j} S_i(1-\alpha)\nu_i\Big)$ ▷ Infection $m \to s$
    $K_i^{s,m} \leftarrow \text{Poisson}\Big(\tau \sum_{j=1}^{A} M(i,j)\beta_s I_{s,j} S_i(1-\alpha)(1-\nu_i)\Big)$ ▷ Infection $s \to m$
    $K_i^{s,s} \leftarrow \text{Poisson}\Big(\tau \sum_{j=1}^{A} M(i,j)\beta_s I_{s,j} S_i(\alpha + (1-\alpha)\nu_i)\Big)$ ▷ Infection $s \to s$
    $K_i^{m} \leftarrow \text{Poisson}(\tau\gamma_m I_{m,i})$ ▷ Number of mild recovery events
    $K_i^{s} \leftarrow \text{Poisson}(\tau\gamma_s I_{s,i})$ ▷ Number of severe recovery events
    **for** $X \in \{m, s\}$ **do**
      **for** $Y \in \{m, s\}$ **do**
        $C_i^{X\to Y} \leftarrow C_i^{X\to Y} + \min(K_i^{X,Y}, S_i)$
        **if** $Y = m$ **then**
          $I_{m,i} \leftarrow I_{m,i} + \min(K_i^{X,Y}, S_i)$
        **else if** $Y = s$ **then**
          $I_{s,i} \leftarrow I_{s,i} + \min(K_i^{X,Y}, S_i)$
        **end if**
        $S_i \leftarrow \max(S_i - K_i^{X,Y}, 0)$
      **end for**
    **end for**
    $I_{m,i} \leftarrow \max(I_{m,i} - K_i^{m}, 0)$
    $R_i \leftarrow R_i + \min(K_i^{m}, I_{m,i})$
    $I_{s,i} \leftarrow \max(I_{s,i} - P_i^{s}, 0)$
    $R_i \leftarrow R_i + \min(K_i^{s}, I_{s,i})$
  **end for**
  $t \leftarrow t + \tau$
**end while**
**Return:** $\mathbf{C}^{m\to m}, \mathbf{C}^{m\to s}, \mathbf{C}^{s\to m}, \mathbf{C}^{s\to s}$

---

the data, the likelihood of $\theta = (\alpha, \nu)$ then obeys

$$\begin{aligned}
L(\theta|\tilde{C}^{m\to m}, \tilde{C}^{m\to s}, \tilde{C}^{s\to m}, \tilde{C}^{s\to s}) \propto\, & \mathbb{P}(\tilde{C}^{m\to m}, \tilde{C}^{m\to s}, \tilde{C}^{s\to m}, \tilde{C}^{s\to s}|\theta) \\
=& \prod_X \prod_Y \mathbb{P}(X \to Y)^{\tilde{C}^{X\to Y}} \\
=& \big(\alpha + (1-\alpha)(1-\nu)\big)^{\tilde{C}^{m\to m}} \\
& \times \big((1-\alpha)\nu\big)^{\tilde{C}^{m\to s}} \\
& \times \big((1-\alpha)(1-\nu)\big)^{\tilde{C}^{s\to m}} \\
& \times \big(\alpha + (1-\alpha)\nu\big)^{\tilde{C}^{s\to s}}
\end{aligned}$$

We denote the parameter values maximising this likelihood (the MLE) by $\hat{\theta} = (\hat{\alpha}, \hat{\nu})$. For computational efficiency and to avoid rounding errors, we worked with the log-likelihood:

$$\hat{\theta} = \underset{\theta}{\operatorname{argmax}} \log(L(\theta|\tilde{C}^{m\to m}, \tilde{C}^{m\to s}, \tilde{C}^{s\to m}, \tilde{C}^{s\to s}))$$

##### Age-dependent likelihood

In the age-dependent setting, we assumed the availability of aggregate contract tracing data that is stratified by secondary case age group. We denote $\tilde{C}_i^{X\to Y}$ as the number of pairs with primary case severity X and secondary case severity Y, where the secondary case is in age group $i$. The likelihood of $\theta = (\alpha, \nu_1, \ldots, \nu_A)$, where $A$ is the number of age groups, then obeys

$$\begin{aligned}
L(\theta|\tilde{\mathbf{C}}^{m\to m}, \tilde{\mathbf{C}}^{m\to s}, \tilde{\mathbf{C}}^{s\to m}, \tilde{\mathbf{C}}^{s\to s}) \propto & \prod_i \mathbb{P}(\tilde{C}_i^{m\to m}, \tilde{C}_i^{m\to s}, \tilde{C}_i^{s\to m}, \tilde{C}_i^{s\to s}|\theta) \\
=& \prod_i \prod_X \prod_Y \mathbb{P}(X \to Y|\text{ secondary case is age } i)^{\tilde{C}_i^{X\to Y}} \\
=& \prod_i \Big( \big(\alpha + (1-\alpha)(1-\nu_i)\big)^{\tilde{C}_i^{m\to m}} \\
& \times \big((1-\alpha)\nu_i\big)^{\tilde{C}_i^{m\to s}} \\
& \times \big((1-\alpha)(1-\nu_i)\big)^{\tilde{C}_i^{s\to m}} \\
& \times \big(\alpha + (1-\alpha)\nu_i\big)^{\tilde{C}_i^{s\to s}} \Big)
\end{aligned}$$

#### S1.5 Applying MCMC

To estimate parameters from age-structured data, we used the Adaptive Metropolis algorithm (Algorithm 3), performing a total of 30,000 iterations with a burn-in period of 3,000 iterations. The posterior distributions are then generated from the remaining 27,000 iterations.

In this case, $\hat{\alpha}$ is defined to be the mean of the marginal posterior distribution for $\alpha$ (and equivalently for $\hat{\nu_i}$), instead of being the MLE.

To ensure that the covariance matrix converged to an optimal value within 30,000 iterations, we varied $\Sigma_0$ according to the number of primary-secondary case pairs, $n$. Since the likelihood, and therefore the acceptance probability, decreases with increasing $n$, a smaller covariance is required to maintain a reasonable acceptance rate. Specifically, we set

$$\Sigma_0 = \frac{1}{n} I_{A+1},$$

where $I_{A+1}$ is the identity matrix with dimension $A+1$ and $A$ is the number of age groups. I initialised $\theta_0$ such that $\alpha_0 = 0.2$ and $\nu_{0,i} = 0.1$ across all age groups.

**Algorithm 3** Adaptive Metropolis algorithm

**Input**: $\pi, D, d$ ▷ Prior distribution, observed data, no. of parameters
**Input**: $N, \Sigma_0, n_0$ ▷ No. of iterations, initial covariance, start of update iteration
**Output**: $\mathcal{S}$ ▷ Accepted samples
$\mathcal{S} = \emptyset$
$\theta_0 \sim \pi(\theta)$ ▷ Set $\theta_0$ from the prior
**for** $n = 1 : N$ **do**
  $\theta' \sim \mathcal{N}(\theta_{n-1}, \frac{2.38^2}{d}\Sigma_{n-1})$ ▷ Propose new parameters
  $\alpha = \min\left\{1, \frac{\mathcal{L}(D|\theta')\pi(\theta')}{\mathcal{L}(D|\theta_{n-1})\pi(\theta_{n-1})}\right\}$ ▷ Calculate the acceptance probability
  $u \sim \text{Unif}[0, 1]$
  **if** u $< \alpha$ **then** ▷ Accept $\theta'$ with probability given by $\alpha$
    $\theta_n = \theta'$
  **else**
    $\theta_n = \theta_{n-1}$
  **end if**
  $S = S \cup \{\theta_n\}$ ▷ Add updated state to sample set
  $\mu_n = \mu_{n-1} + \frac{1}{n}(\theta_n - \mu_{n-1})$ ▷ Update mean and covariance matrix
  **if** $n \leq n_0$ **then**
    $\Sigma_n = \Sigma_0$
  **else**
    $\Sigma_n = \Sigma_{n-1} + \frac{1}{n}\{(\theta_n - \mu_{n-1})(\theta_n - \mu_{n-1})^T - \Sigma_{n-1}\}$
  **end if**
**end for**

Switching to an MCMC algorithm means careful attention is needed on how values for the strength of symptom propagation, $\alpha$ are proposed. If proposed parameter sets are generated using a multivariate normal distribution, the probability that we propose exactly $\alpha = 0$ is zero. Therefore, if we restrict $\alpha \geq 0$, the mean of our posterior distribution will never be zero, regardless of the underlying data. Furthermore, $\alpha = 0$ will never be contained within the 95% credible interval.

One solution to this is to introduce a slab-and-spike prior [45] on $\alpha$, which explicitly allocates a positive probability mass at $\alpha = 0$. Under this approach, the prior is formulated as a mixture between a point mass at zero (the "spike") and a continuous distribution over $(0, 1]$ (the "slab"). During MCMC, proposals for $\alpha$ are therefore drawn either from the slab component or from the spike component (i.e. $\alpha = 0$). This ensures that the posterior distribution can place mass at $\alpha = 0$ when the data is consistent with there being no symptom propagation.

However, an alternative solution is to allow for the consideration of $\alpha < 0$ (corresponding to severe infections being more likely to lead to mild infections and vice versa). In our view, this is a more natural approach since, from a mathematical perspective, postive and negative $\alpha$ are equally feasible.

Specifically, we can think of $\alpha$ as the increase in the probability of having severe symptoms when infected by a severe infector, versus being infected by someone with mild symptoms. However, it is theoretically possible that you would be more likely to have severe symptoms if infected by someone with mild symptoms (and vice versa). This scenario corresponds to $\alpha < 0$. Indeed, when $\alpha = 0$, we are equally likely to have $C^{m\to m} + C^{s\to s} > C^{s\to m} + C^{m\to s}$ (corresponding to $\hat{\alpha} > 0$) as we are to have $C^{m\to m} + C^{s\to s} < C^{s\to m} + C^{m\to s}$ (corresponding to $\hat{\alpha} < 0$).

When considering $\alpha < 0$, we have to be careful to ensure that our probabilities remain in $[0, 1]$. That validity condition requires these four criteria are satisfied:

$$\begin{aligned} 0 \leq \alpha + (1-\alpha)(1-\nu) \leq 1, &\qquad 0 \leq (1-\alpha)(1-\nu) \leq 1, \\ 0 \leq \alpha + (1-\alpha)\nu \leq 1, &\qquad 0 \leq (1-\alpha)\nu \leq 1. \end{aligned}$$

First, note that, since $\alpha < 0$, our requirements simplify to,

$$\begin{aligned} 0 \leq \alpha + (1-\alpha)(1-\nu), &\qquad (1-\alpha)(1-\nu) \leq 1, \\ 0 \leq \alpha + (1-\alpha)\nu, &\qquad (1-\alpha)\nu \leq 1, \end{aligned}$$

which hold if

$$\alpha \geq -\max\left(\frac{1-\nu}{\nu}, \frac{\nu}{1-\nu}\right).$$

Therefore, we chose my priors to be uniform for $\nu_i \in [0, 1]$ and $\alpha \in \left[-\max_i\left(\frac{1-\nu_i}{\nu_i}, \frac{\nu_i}{1-\nu_i}\right), 1\right]$, and zero outside of these ranges. We chose these priors to simulate a situation where we have no previous knowledge relating to the symptom propagation parameters (beyond which values are mathematically feasible), as might occur for a novel pathogen of public health concern.

Throughout this analysis, randomly selected trace plots were analysed to ensure the chains were well-mixed and converged. Across all replicates, acceptance rates were monitored to ensure they remained within a reasonable range (0.15–0.30).

### S2 Additional age-free results

#### S2.1 Errors in $\nu$

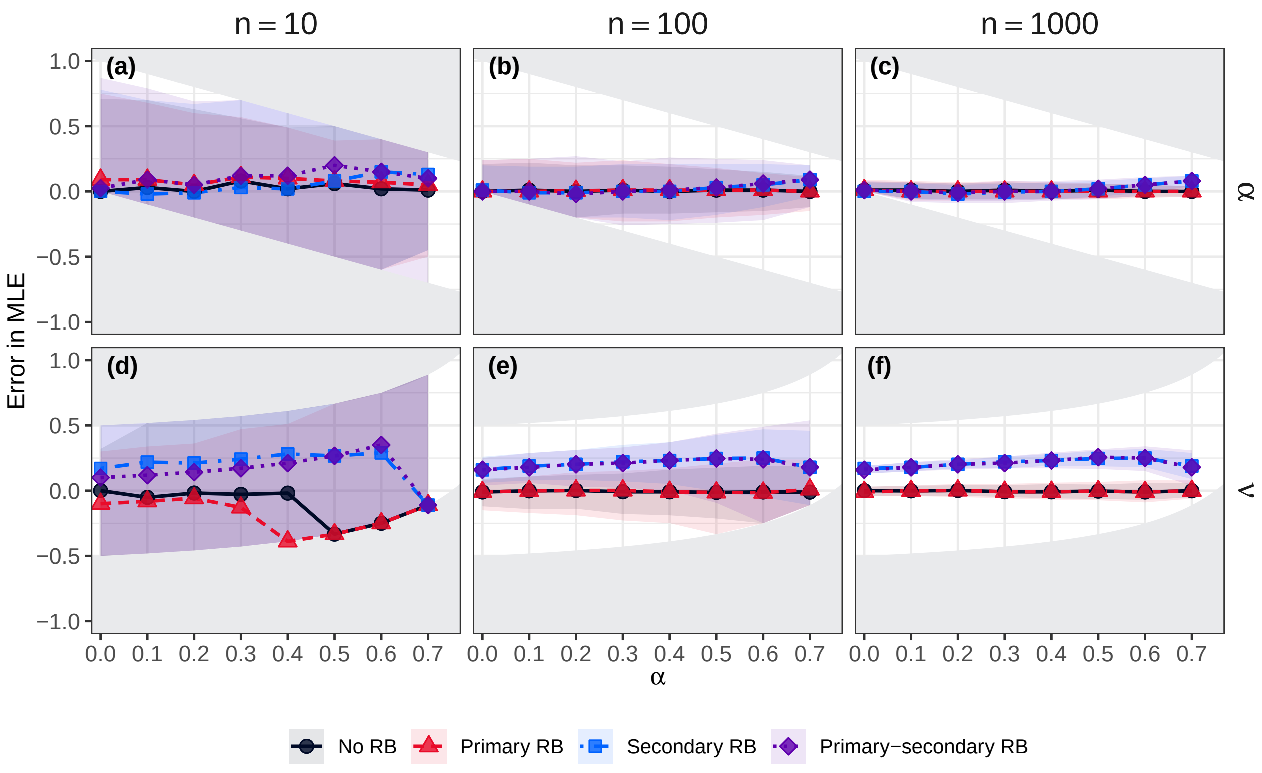

**Fig. S1. Error in the predicted values of $\hat{\alpha}$ and $\hat{\nu}$.** The error in the predicted values of $\hat{\alpha}$ (top row) and $\hat{\nu}$ (bottom row), given by the maximum likelihood estimate, for four reporting bias scenarios: no RB (black circles), primary case RB (red triangles), secondary case RB (blue squares) and primary and secondary case RB (purple diamonds). A positive error corresponds to the predicted value being an overestimate of the true value; a negative error corresponds to the predicted value being an underestimate of the true value. The solid grey regions correspond to infeasible errors, which are present due to the restriction of $\hat{\alpha}$ and $\hat{\nu}$ being between 0 and 1. The points show the median errors, and the dashed lines show the interpolation between those points. The shaded area spans the 2.5th percentile to the 97.5th percentile (the 95% uncertainty interval). Across panels, the assessed sample sizes (i.e. the number of data points used in each replicate) vary. Each column corresponds to the number of contact tracing data points: **(a, d)** 10, **(b, e)** 100, **(c, f)** 1,000.

### S3 Additional age-dependent results

#### S3.1 Motivation

As motivation for the age-dependent analysis performed in this paper, we first applied the age-free methodology to synthetic data generated with an underlying age structure, i.e. age-dependent mixing and age-dependent values of the baseline probability of severe disease, $\nu$.

The addition of age structure in the synthetic data generation process results led to the previous age-free methodological approach overestimating the strength of symptom propagation and finding increased support for the presence of symptom propagation (Fig. S2). Given symptom propagation being absent (i.e. true $\alpha = 0$), the median error increased to as high as 0.09 and 68% of replicates supported symptom propagation being present (when $n = 1,000$, Fig. S2(c) and (f)).

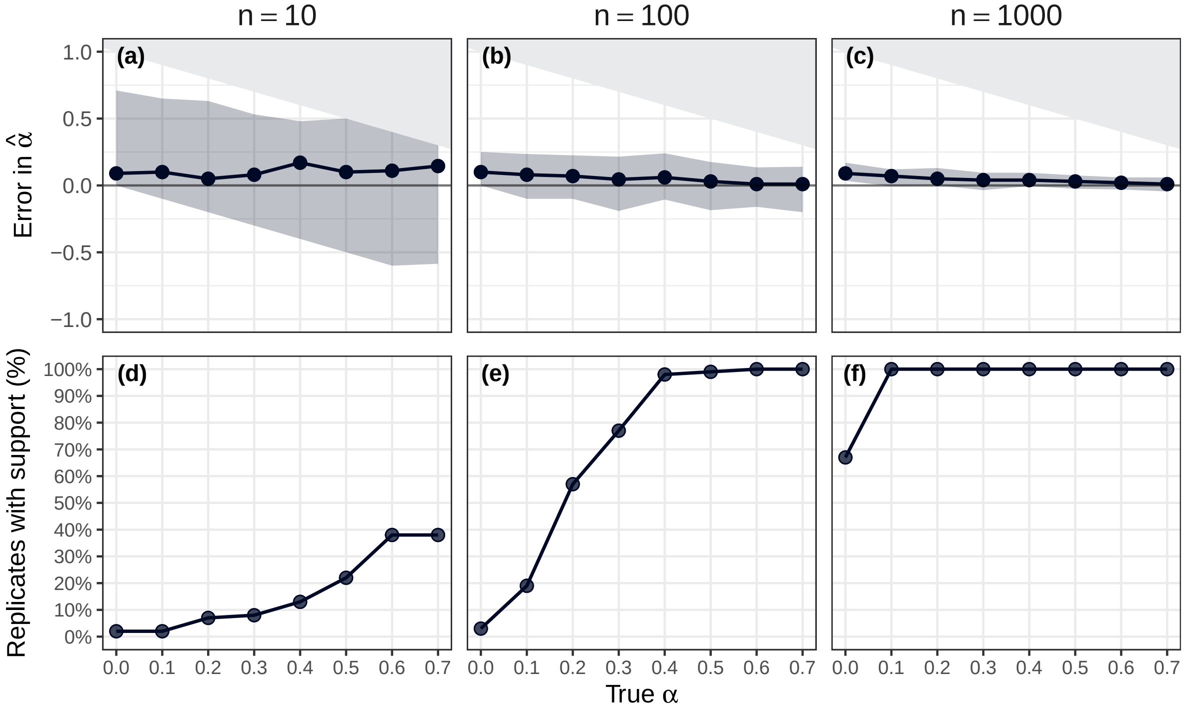

**Fig. S2. Error in the estimated values of $\hat{\alpha}$ and the percentage of replicates with support for symptom propagation, against the true values of $\alpha$: Age-free methodology.** The top row shows the error in the estimates of $\alpha$. A positive error corresponds to the estimated value being an overestimate of the true value. The solid grey regions correspond to infeasible errors, which are present due to the restriction of $\hat{\alpha}$ being between 0 and 1. The points show the median errors and the lines show the interpolation between those points. The shaded area spans the 2.5th percentile to the 97.5th percentile (the 95% uncertainty interval). The bottom row gives the percentage of replicates (out of 1000) for which $\alpha = 0$ was not within the 95% confidence region (i.e. support for symptom propagation). Across panels, the assessed sample sizes $N_{CT}$ (i.e. the number of data points used in each replicate) vary. Each column corresponds to the number of contact tracing data points: **(a, d)** 10, **(b, e)** 100, **(c, f)** 1,000.

#### S3.2 Posterior distributions

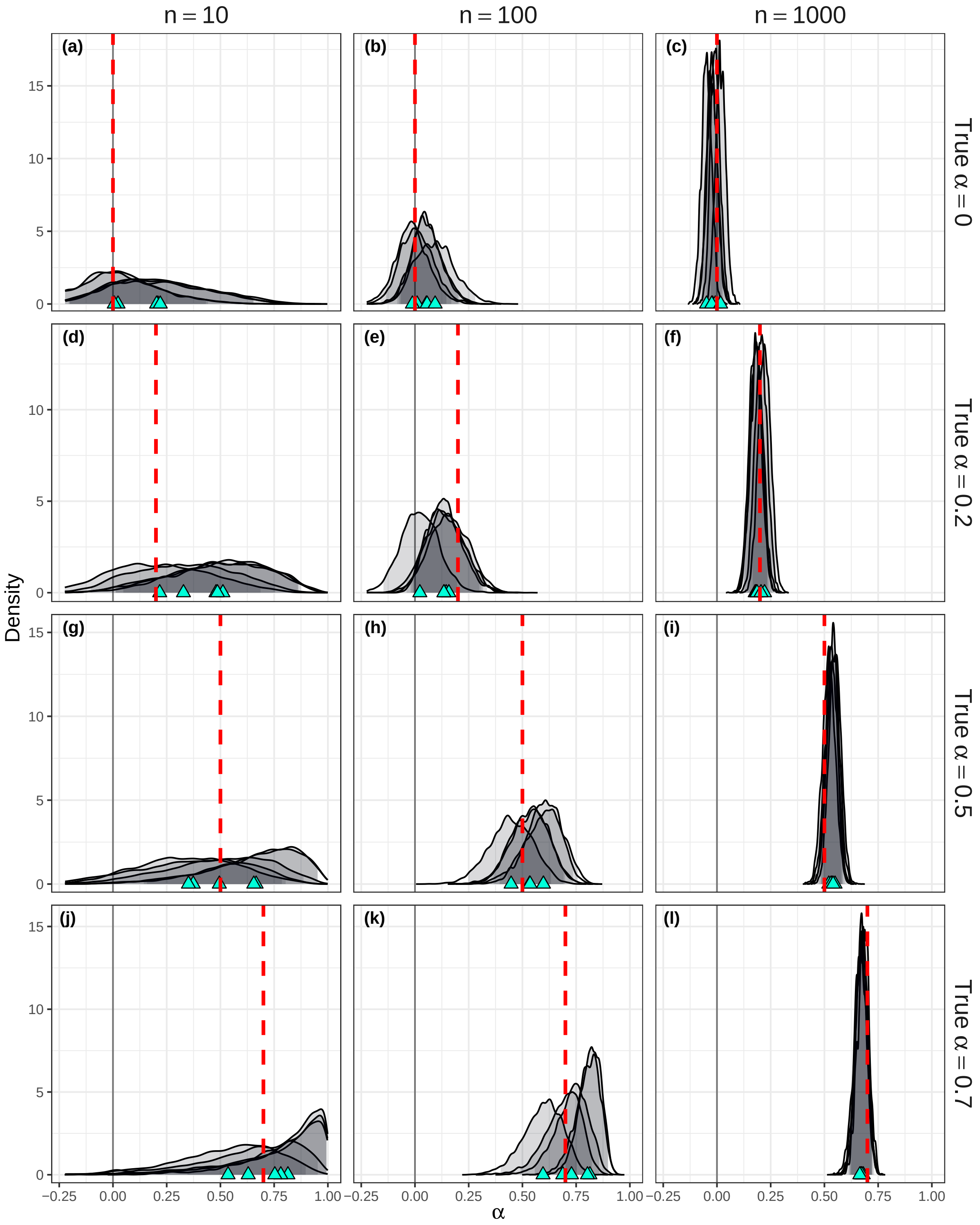

**Fig. S3. Posterior distributions for $\alpha$, across four true $\alpha$ values and three values of $N_{CT}$.** The grey shaded regions indicate the 95% credible intervals for each replicate, and the cyan triangles indicate the posterior means. The grey solid line marks $\alpha = 0$. The red dotted lines correspond to the true values being estimated, which varied across rows: **(a-c)** $\alpha = 0$, **(d-f)** $\alpha = 0.2$, **(g-i)** $\alpha = 0.5$, **(j-l)** $\alpha = 0.7$. Across columns, the number of contact tracing data points varies: **(a, d, g, i)** 10, **(b, e, h, k)** 100, **(c, f, i, l)** 1,000.

#### S3.3 MCMC applied to data with no age-structure

Even when the data has no underlying age structure, we verified that using MCMC leads to slight positive errors for small true $\alpha$ and slight negative errors for larger true $\alpha$ when $N_{CT} = 10$ (Fig. S4).

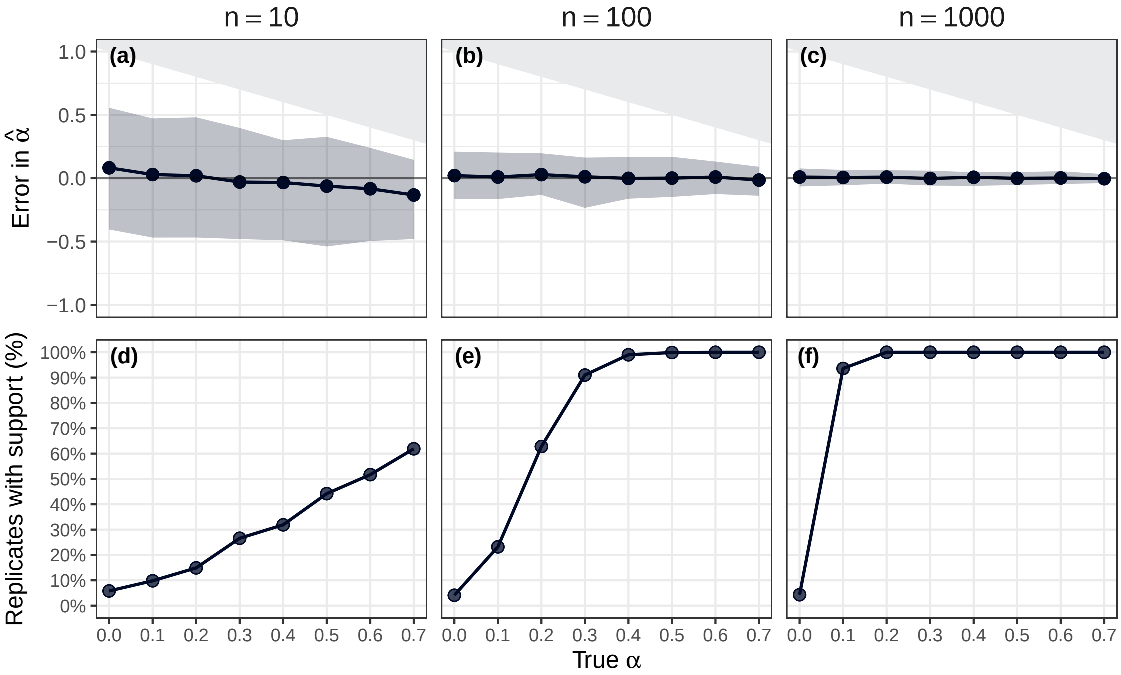

**Fig. S4.** Errors in $\hat{\alpha}$ using MCMC with data generated with no age structure and an age-free likelihood

#### S3.4 Accuracy of $\nu$ estimates

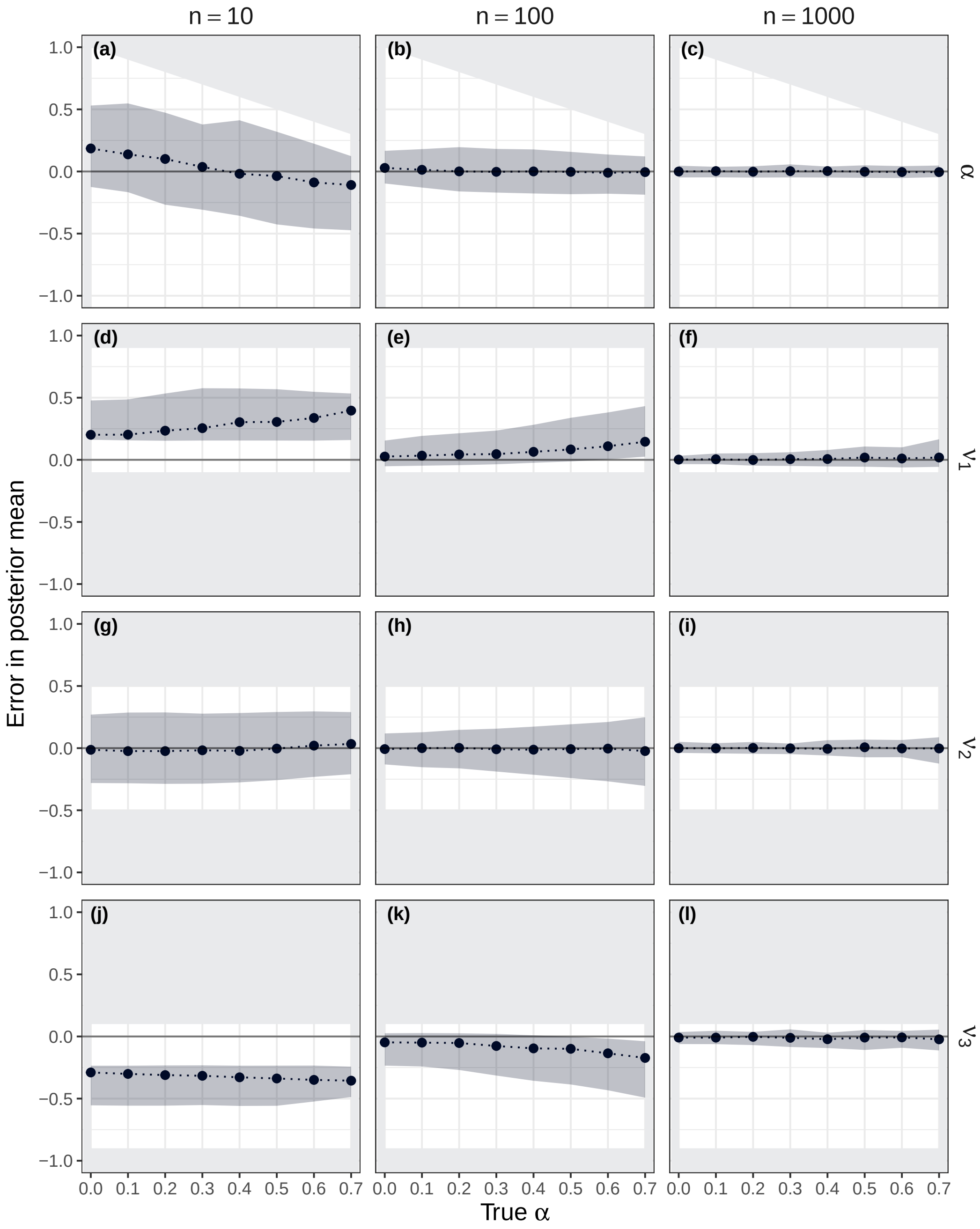

**Fig. S5. Error in the predicted values of $\hat{\alpha}$ and three age-dependent $\hat{\nu}$ values across varying true $\alpha$: realistic age structure.** The error in the predicted values of $\hat{\alpha}$ and $\hat{\nu_1}, \hat{\nu_2}, \hat{\nu_3}$, given by the posterior means. A positive error corresponds to the predicted value being an overestimate of the true value; a negative error corresponds to the predicted value being an underestimate of the true value. The solid grey regions correspond to infeasible errors, which are present due to the restriction of $\hat{\alpha}$ being less than 1 and $\nu_i$ between 0 and 1. The points show the median errors, and the dashed lines show the interpolation between those points. The shaded area spans the 2.5th percentile to the 97.5th percentile (the 95% uncertainty interval). Across columns, the number of contact tracing data points varies: **(a, d, g, i)** 10, **(b, e, h, k)** 100, **(c, f, i, l)** 1000.

### S4 Robustness analysis

#### S4.1 Age-free

We explored the robustness of our age-free results to our parameter choices. In each robustness analysis we varied one of the following from the default parameter value, with all other model parameters remaining unchanged: the relative transmissibility of severe cases ($b = 0.5, 1, 4$), the proportion of cases that were severe ($\mathrm{P} = 0.2, 0.8$) or $R_0$ ($= 1.5, 10$). We also considered the effect of stronger reporting bias. Specifically, severe cases were four times as likely to be reported as mild cases (compared to two in the main results). We generally observed similar results to our main analysis using default values for all model parameters.

Firstly, we considered the error in $\hat{\alpha}$. In the no reporting bias scenario, the median errors remained close to zero. However, the positive error seen in the main text for secondary and primary-secondary reporting bias and larger true $\alpha$ was more pronounced for certain alternate parametrisations ($b = 4$, Fig. S8; $P = 0.2$, Fig. S11). An additional difference was that in some cases secondary or primary-secondary reporting bias led to a negative error in $\hat{\alpha}$, i.e. the strength of symptom propagation was underestimated, for some parameter values ($b = 0.5$, Fig. S6; $\mathrm{P} = 0.8$, Fig. S12), whereas in the main results the errors were either essentially zero, or slightly positive.

We also considered the effect of stronger reporting bias. Specifically, severe cases were four times as likely to be reported as mild cases (compared to two in the main results). Similarly, we found that for some true values of $\alpha$, secondary or primary-secondary reporting bias led to a negative error in $\hat{\alpha}$ Our perspective is that this is not a concern, as underestimating symptom propagation would only lead to more conservative estimates, particularly given the standard assumption is that symptom propagation does not occur.

Varying our parameter choices had little effect on our general qualitative findings for the support for symptom propagation. However, we did find a noticeable reduction in support for symptom propagation for $N_{CT} = 10$ (compared to the main text results) for some parameters, namely $b = 4$ (Fig. S8) and $\mathrm{P} = 0.8$ (Fig. S12). This effect was generally not seen for larger values of $N_{CT}$.

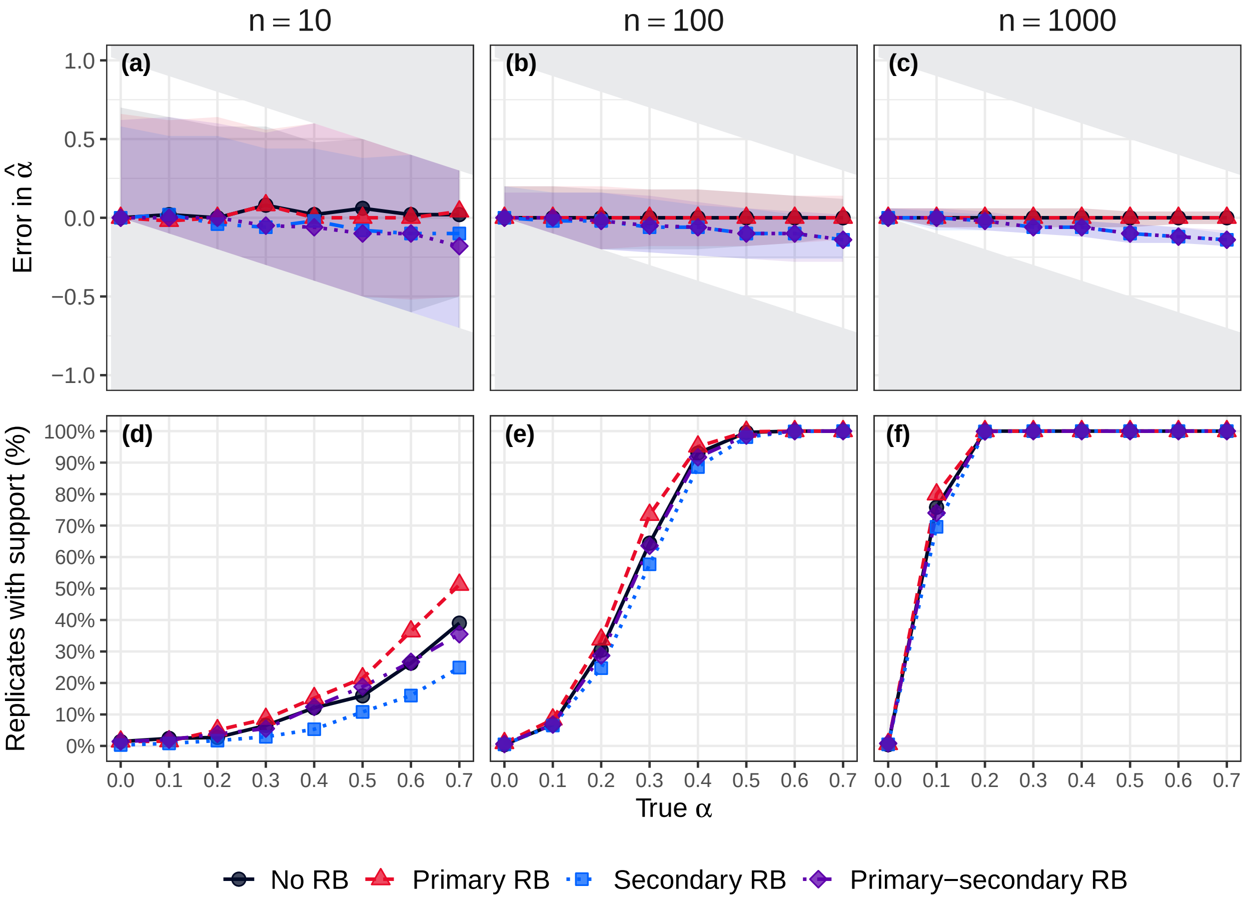

**Fig. S6. Error in the estimated values of $\hat{\alpha}$ and the percentage of replicates with support for symptom propagation: $\mathbf{b = 0.5}$** We consider four reporting bias scenarios: no reporting bias (black circles), primary case reporting bias (red triangles), secondary case reporting bias (blue squares) and primary and secondary case reporting bias (purple diamonds). The top row shows the error in the estimates of $\hat{\alpha}$ . A positive error corresponds to the estimated value being an overestimate of the true value of $\alpha$. The solid grey regions correspond to infeasible errors, which are present due to the restriction of $\hat{\alpha}$ being between 0 and 1. The points show the median errors and the lines show the interpolation between those points. The shaded area spans the 2.5th percentile to the 97.5th percentile (the 95% uncertainty interval). The bottom row gives the percentage of replicates (out of 1000) for which $\alpha = 0$ was not within the 95% confidence region (i.e. support for symptom propagation). Across panels, we vary the assessed sample sizes $N_{CT}$ (i.e. the number of data points used in each replicate). Each column corresponds to the number of contact tracing data points: **(a, d)** 10, **(b, e)** 100, **(c, f)** 1,000.

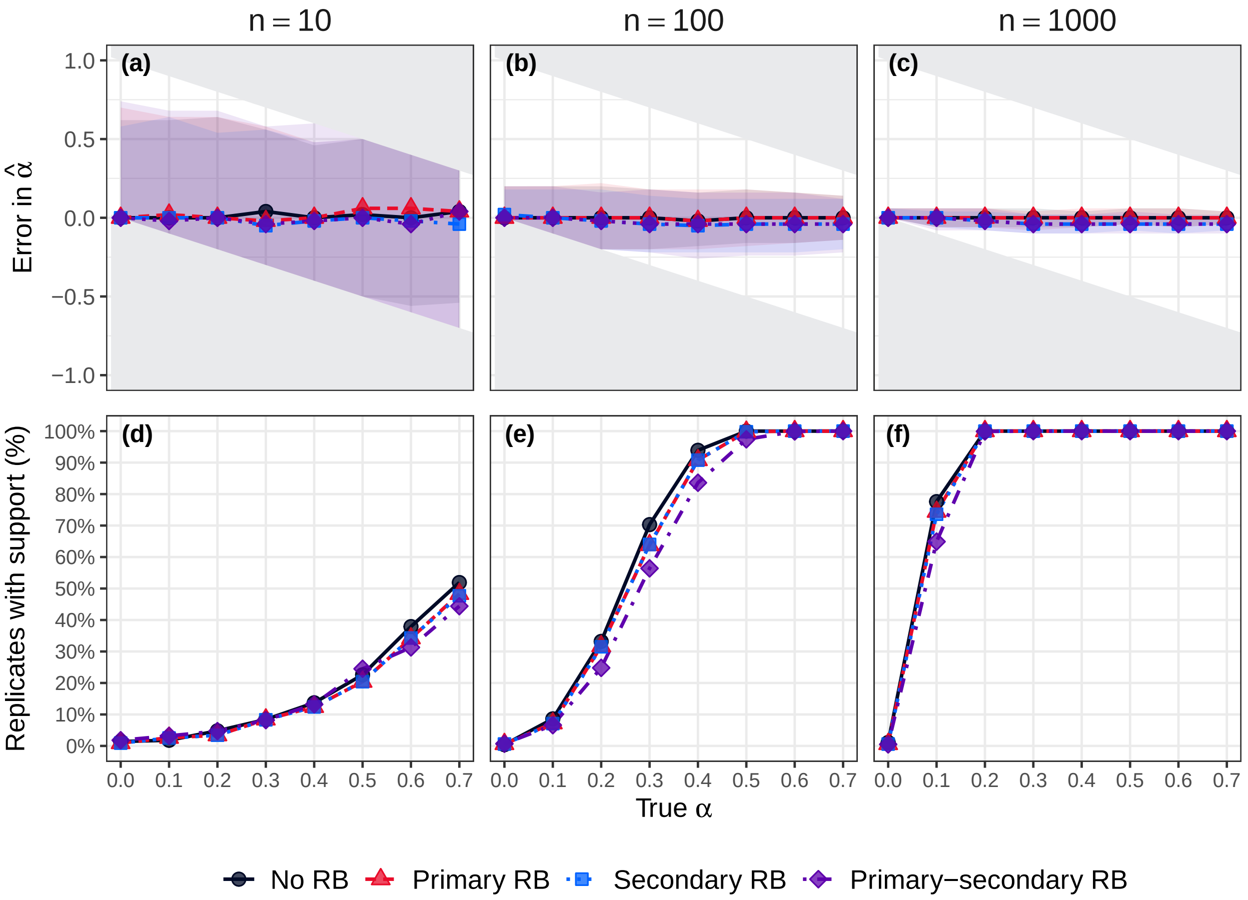

**Fig. S7. Error in the estimated values of $\hat{\alpha}$ and the percentage of replicates with support for symptom propagation: b = 1** We consider four reporting bias scenarios: no reporting bias (black circles), primary case reporting bias (red triangles), secondary case reporting bias (blue squares) and primary and secondary case reporting bias (purple diamonds). The top row shows the error in the estimates of $\hat{\alpha}$ . A positive error corresponds to the estimated value being an overestimate of the true value of $\alpha$. The solid grey regions correspond to infeasible errors, which are present due to the restriction of $\hat{\alpha}$ being between 0 and 1. The points show the median errors and the lines show the interpolation between those points. The shaded area spans the 2.5th percentile to the 97.5th percentile (the 95% uncertainty interval). The bottom row gives the percentage of replicates (out of 1000) for which $\alpha = 0$ was not within the 95% confidence region (i.e. support for symptom propagation). Across panels, we vary the assessed sample sizes $N_{CT}$ (i.e. the number of data points used in each replicate). Each column corresponds to the number of contact tracing data points: **(a, d)** 10, **(b, e)** 100, **(c, f)** 1,000.

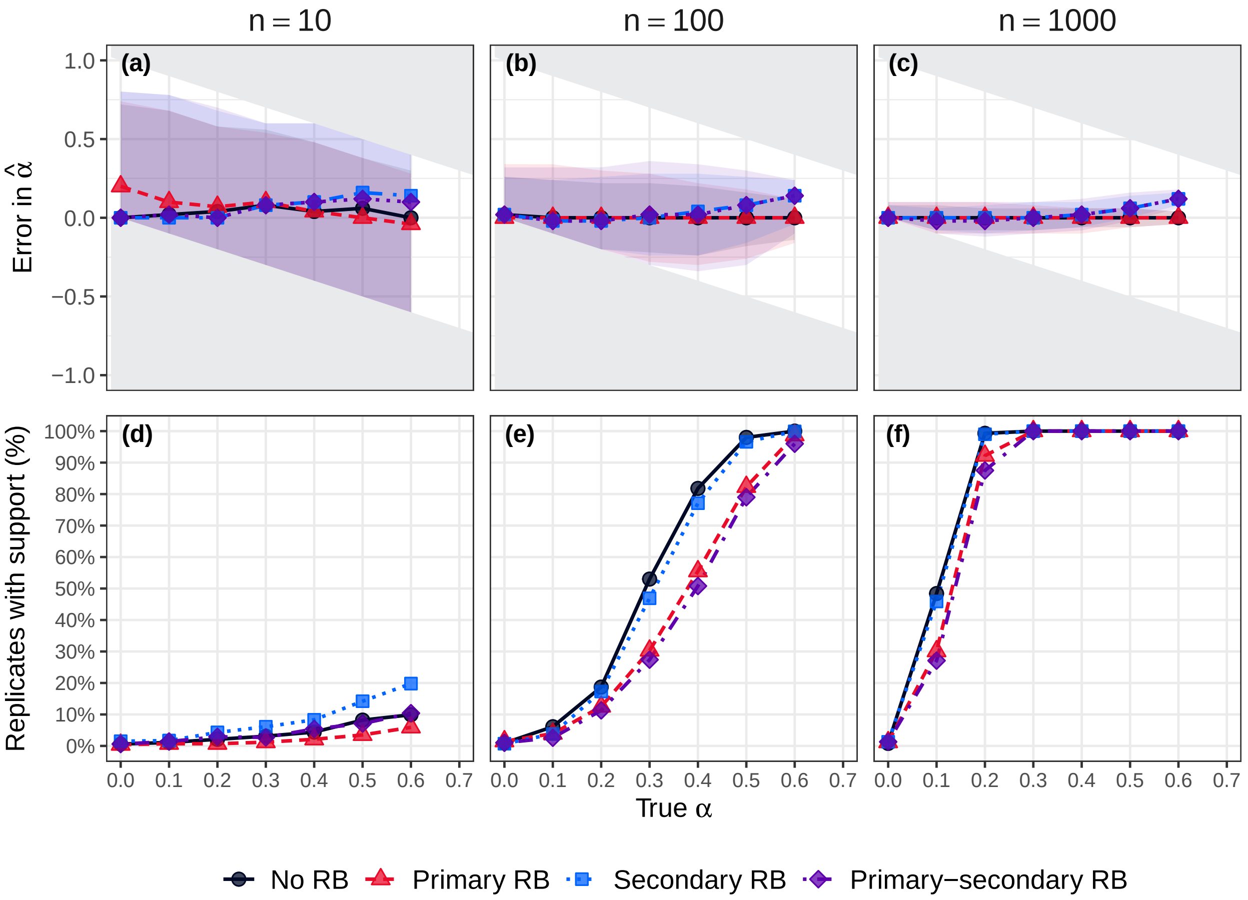

**Fig. S8. Error in the estimated values of $\hat{\alpha}$ and the percentage of replicates with support for symptom propagation: b = 4** We consider four reporting bias scenarios: no reporting bias (black circles), primary case reporting bias (red triangles), secondary case reporting bias (blue squares) and primary and secondary case reporting bias (purple diamonds). The top row shows the error in the estimates of $\hat{\alpha}$ . A positive error corresponds to the estimated value being an overestimate of the true value of $\alpha$. The solid grey regions correspond to infeasible errors, which are present due to the restriction of $\hat{\alpha}$ being between 0 and 1. The points show the median errors and the lines show the interpolation between those points. The shaded area spans the 2.5th percentile to the 97.5th percentile (the 95% uncertainty interval). The bottom row gives the percentage of replicates (out of 1000) for which $\alpha = 0$ was not within the 95% confidence region (i.e. support for symptom propagation). Across panels, we vary the assessed sample sizes $N_{CT}$ (i.e. the number of data points used in each replicate). Each column corresponds to the number of contact tracing data points: **(a, d)** 10, **(b, e)** 100, **(c, f)** 1,000.

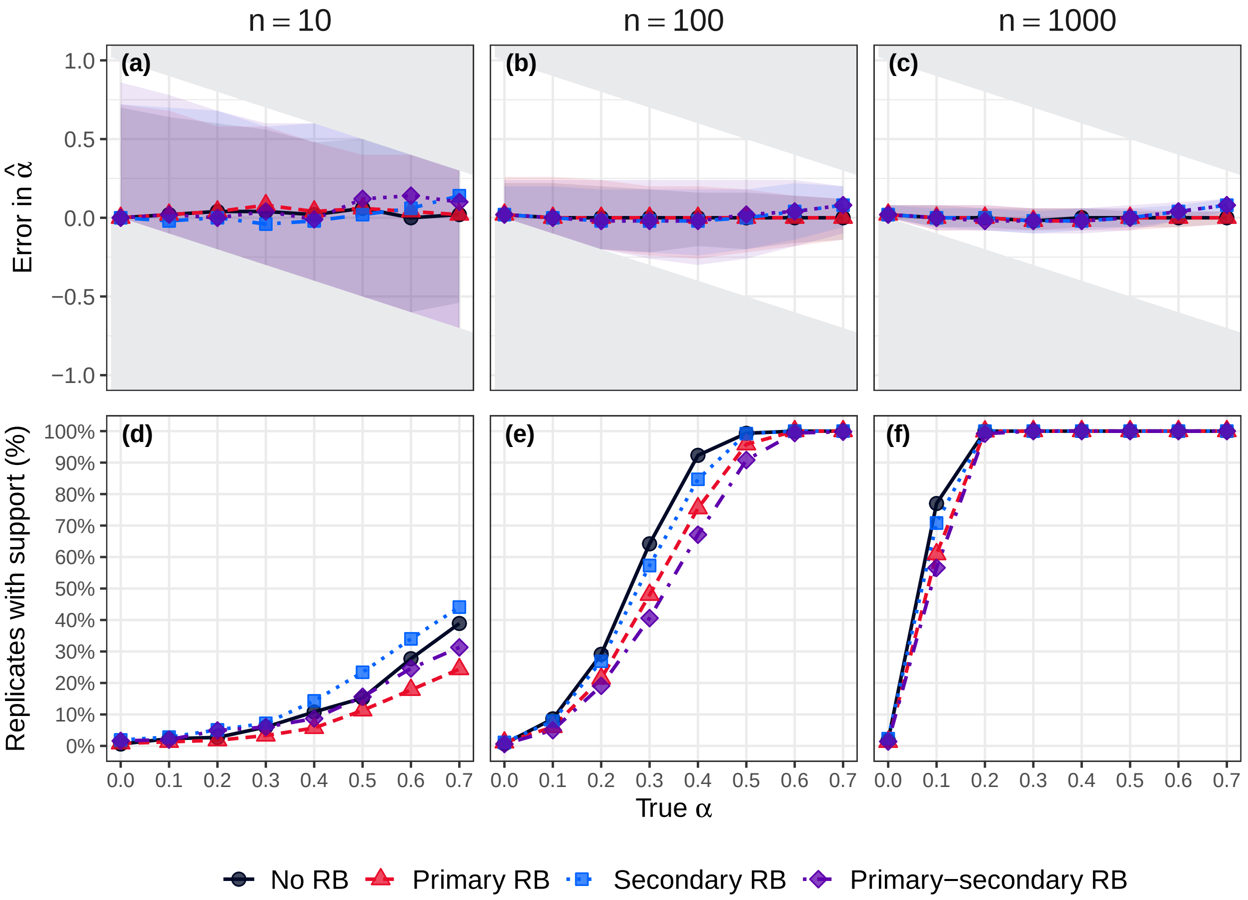

**Fig. S9. Error in the estimated values of $\hat{\alpha}$ and the percentage of replicates with support for symptom propagation: $\mathbf{R_0 = 1.5}$** We consider four reporting bias scenarios: no reporting bias (black circles), primary case reporting bias (red triangles), secondary case reporting bias (blue squares) and primary and secondary case reporting bias (purple diamonds). The top row shows the error in the estimates of $\hat{\alpha}$ . A positive error corresponds to the estimated value being an overestimate of the true value of $\alpha$. The solid grey regions correspond to infeasible errors, which are present due to the restriction of $\hat{\alpha}$ being between 0 and 1. The points show the median errors and the lines show the interpolation between those points. The shaded area spans the 2.5th percentile to the 97.5th percentile (the 95% uncertainty interval). The bottom row gives the percentage of replicates (out of 1000) for which $\alpha = 0$ was not within the 95% confidence region (i.e. support for symptom propagation). Across panels, we vary the assessed sample sizes $N_{CT}$ (i.e. the number of data points used in each replicate). Each column corresponds to the number of contact tracing data points: **(a, d)** 10, **(b, e)** 100, **(c, f)** 1,000.

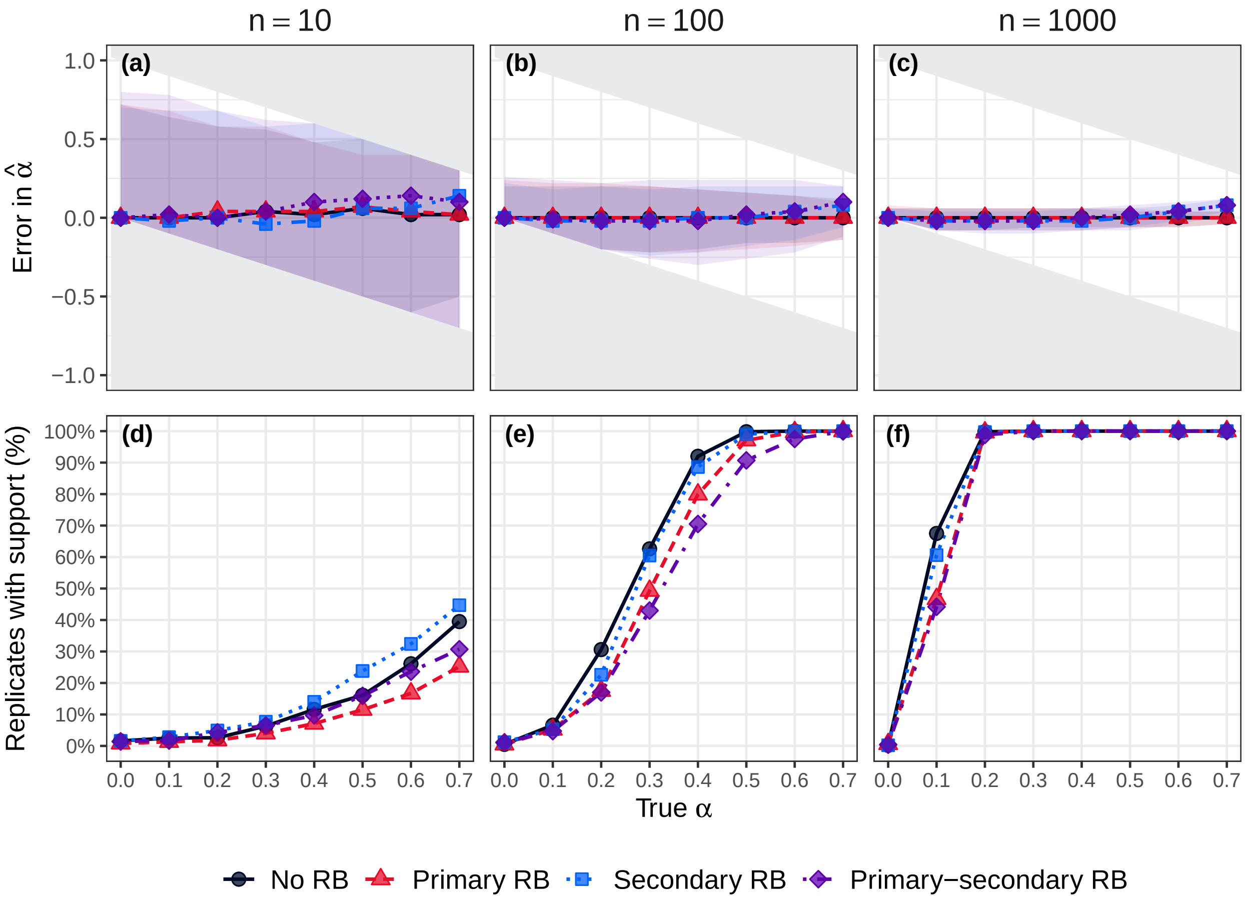

**Fig. S10. Error in the estimated values of $\hat{\alpha}$ and the percentage of replicates with support for symptom propagation: $\mathbf{R_0 = 10}$** We consider four reporting bias scenarios: no reporting bias (black circles), primary case reporting bias (red triangles), secondary case reporting bias (blue squares) and primary and secondary case reporting bias (purple diamonds). The top row shows the error in the estimates of $\hat{\alpha}$ . A positive error corresponds to the estimated value being an overestimate of the true value of $\alpha$. The solid grey regions correspond to infeasible errors, which are present due to the restriction of $\hat{\alpha}$ being between 0 and 1. The points show the median errors and the lines show the interpolation between those points. The shaded area spans the 2.5th percentile to the 97.5th percentile (the 95% uncertainty interval). The bottom row gives the percentage of replicates (out of 1000) for which $\alpha = 0$ was not within the 95% confidence region (i.e. support for symptom propagation). Across panels, we vary the assessed sample sizes $N_{CT}$ (i.e. the number of data points used in each replicate). Each column corresponds to the number of contact tracing data points: **(a, d)** 10, **(b, e)** 100, **(c, f)** 1,000.

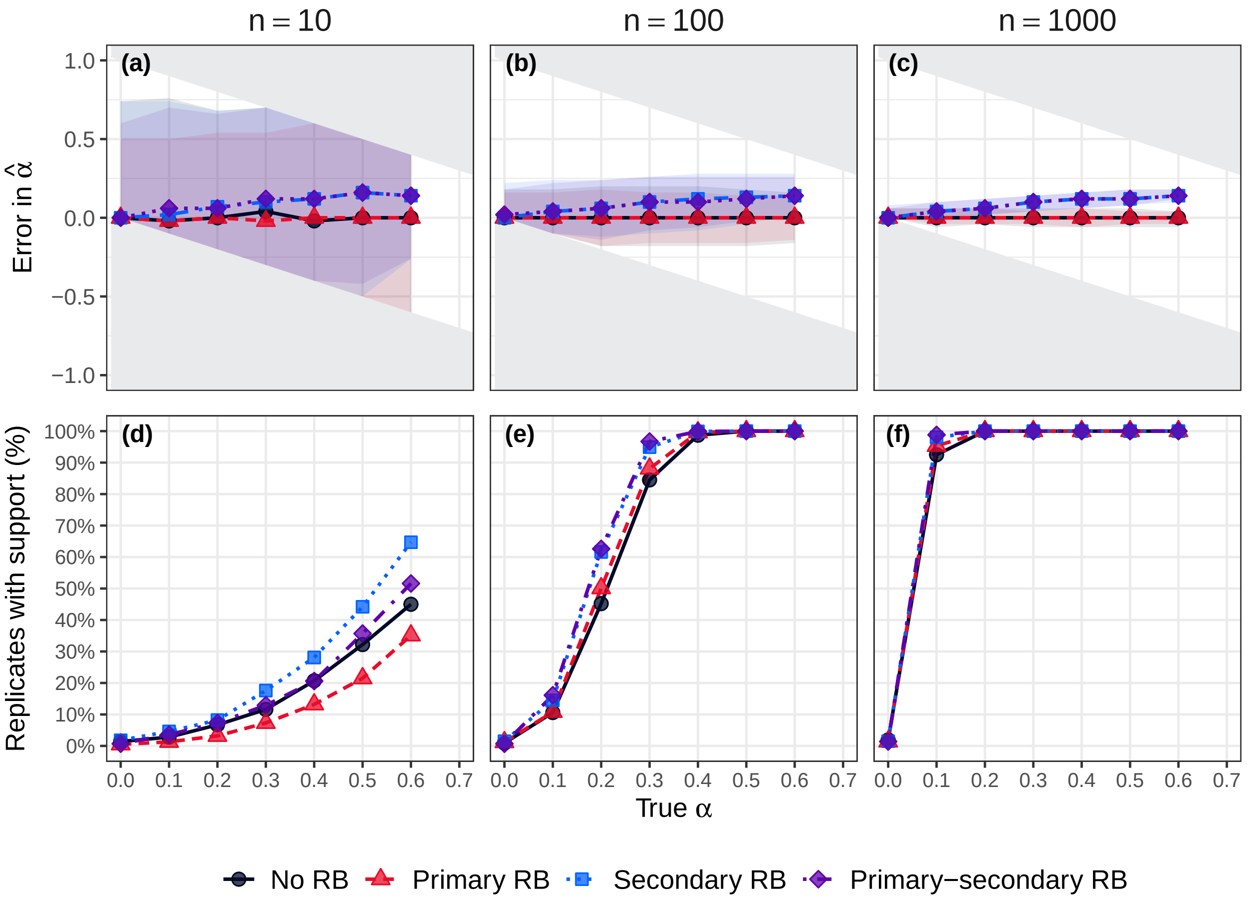

**Fig. S11. Error in the estimated values of $\hat{\alpha}$ and the percentage of replicates with support for symptom propagation: $\mathbf{P = 0.2}$** We consider four reporting bias scenarios: no reporting bias (black circles), primary case reporting bias (red triangles), secondary case reporting bias (blue squares) and primary and secondary case reporting bias (purple diamonds). The top row shows the error in the estimates of $\hat{\alpha}$ . A positive error corresponds to the estimated value being an overestimate of the true value of $\alpha$. The solid grey regions correspond to infeasible errors, which are present due to the restriction of $\hat{\alpha}$ being between 0 and 1. The points show the median errors and the lines show the interpolation between those points. The shaded area spans the 2.5th percentile to the 97.5th percentile (the 95% uncertainty interval). The bottom row gives the percentage of replicates (out of 1000) for which $\alpha = 0$ was not within the 95% confidence region (i.e. support for symptom propagation). Across panels, we vary the assessed sample sizes $N_{CT}$ (i.e. the number of data points used in each replicate). Each column corresponds to the number of contact tracing data points: **(a, d)** 10, **(b, e)** 100, **(c, f)** 1,000.

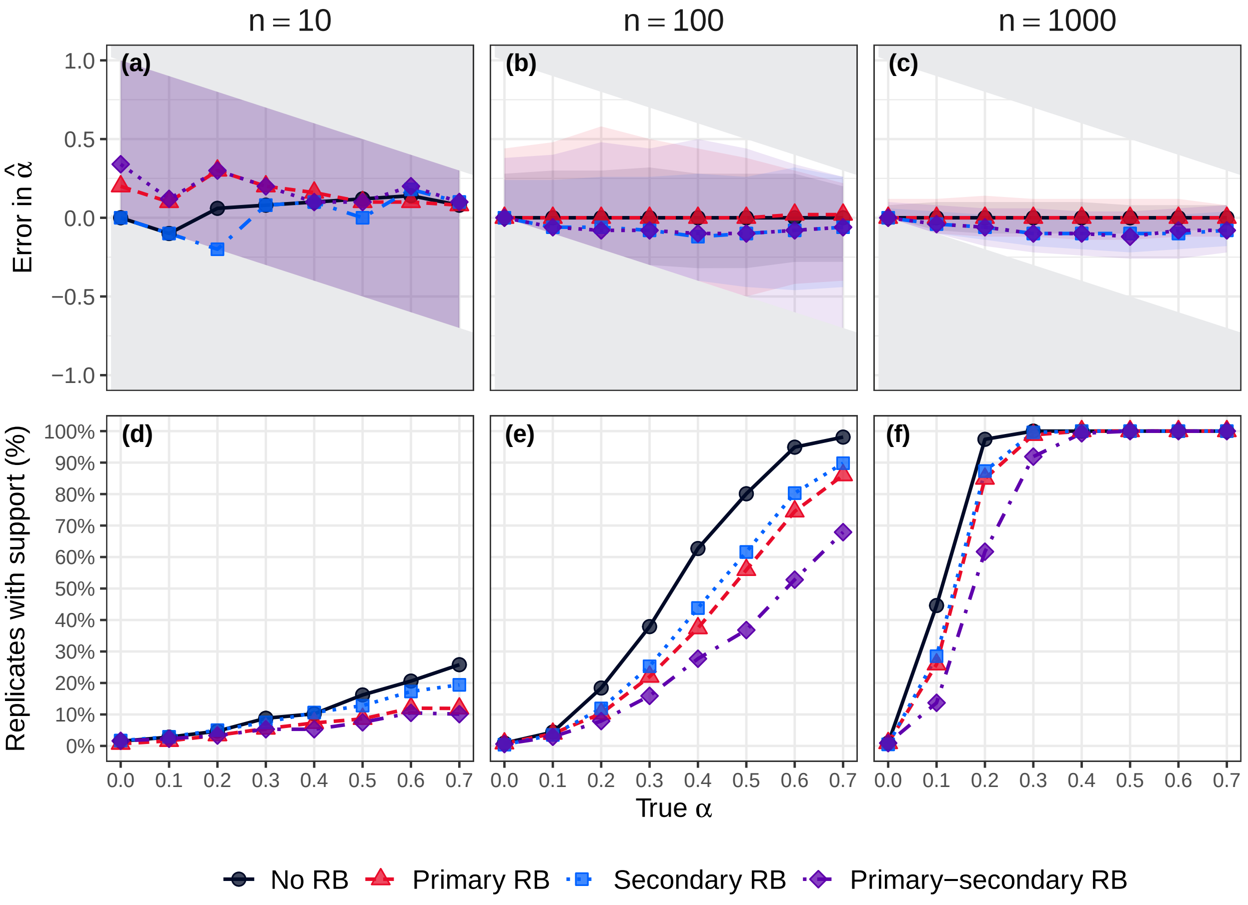

**Fig. S12. Error in the estimated values of $\hat{\alpha}$ and the percentage of replicates with support for symptom propagation: $\mathbf{P = 0.8}$** We consider four reporting bias scenarios: no reporting bias (black circles), primary case reporting bias (red triangles), secondary case reporting bias (blue squares) and primary and secondary case reporting bias (purple diamonds). The top row shows the error in the estimates of $\hat{\alpha}$ . A positive error corresponds to the estimated value being an overestimate of the true value of $\alpha$. The solid grey regions correspond to infeasible errors, which are present due to the restriction of $\hat{\alpha}$ being between 0 and 1. The points show the median errors and the lines show the interpolation between those points. The shaded area spans the 2.5th percentile to the 97.5th percentile (the 95% uncertainty interval). The bottom row gives the percentage of replicates (out of 1000) for which $\alpha = 0$ was not within the 95% confidence region (i.e. support for symptom propagation). Across panels, we vary the assessed sample sizes $N_{CT}$ (i.e. the number of data points used in each replicate). Each column corresponds to the number of contact tracing data points: **(a, d)** 10, **(b, e)** 100, **(c, f)** 1,000.

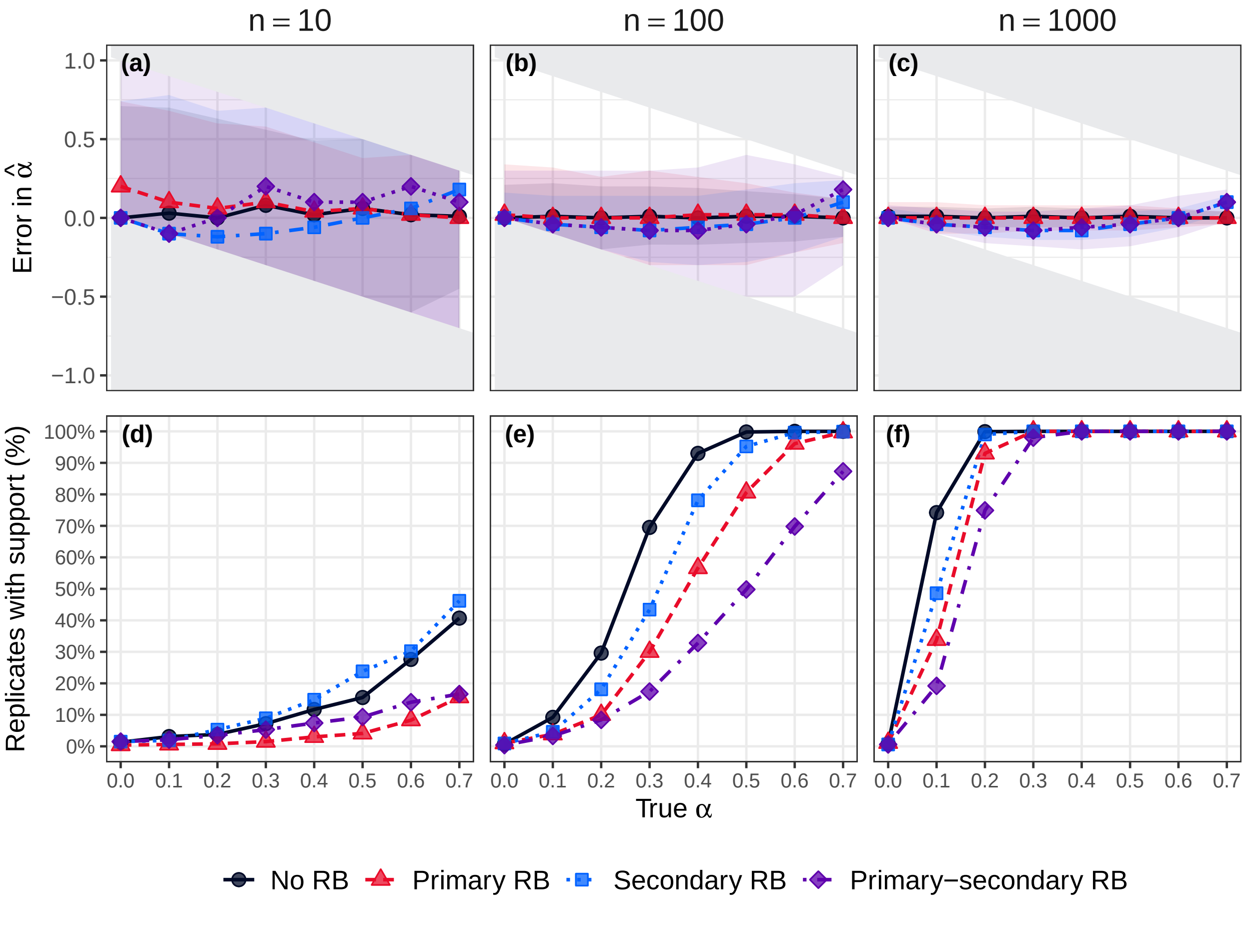

**Fig. S13. Error in the estimated values of $\hat{\alpha}$ and the percentage of replicates with support for symptom propagation: stronger reporting bias** We consider four reporting bias scenarios: no reporting bias (black circles), primary case reporting bias (red triangles), secondary case reporting bias (blue squares) and primary and secondary case reporting bias (purple diamonds). The top row shows the error in the estimates of $\hat{\alpha}$ . A positive error corresponds to the estimated value being an overestimate of the true value of $\alpha$. The solid grey regions correspond to infeasible errors, which are present due to the restriction of $\hat{\alpha}$ being between 0 and 1. The points show the median errors and the lines show the interpolation between those points. The shaded area spans the 2.5th percentile to the 97.5th percentile (the 95% uncertainty interval). The bottom row gives the percentage of replicates (out of 1000) for which $\alpha = 0$ was not within the 95% confidence region (i.e. support for symptom propagation). Across panels, we vary the assessed sample sizes $N_{CT}$ (i.e. the number of data points used in each replicate). Each column corresponds to the number of contact tracing data points: **(a, d)** 10, **(b, e)** 100, **(c, f)** 1,000.

##### S4.2 Age-dependent

We also considered the effect of varying the transmission rates for the age-dependent analysis from a baseline scenario of $b = 2$, $\beta = 0.25$ (where $\beta_M = \beta$, $\beta_S = b\beta$). We first considered severe cases being even more transmissible than mild cases ($b = 4$, $\beta = 0.25$; Fig. S14). Then, we considered severe cases being equally transmissible ($b = 1$; Fig. S15) or less transmissible ($b = 0.5$; Fig. S16) than mild cases. In these cases, we set $\beta = 0.5$ to account for the overall reduction in severe from making these changes. Finally, we considered keeping $b = 2$, but increasing the transmissibility to $\beta = 0.5$ (Fig. S17).

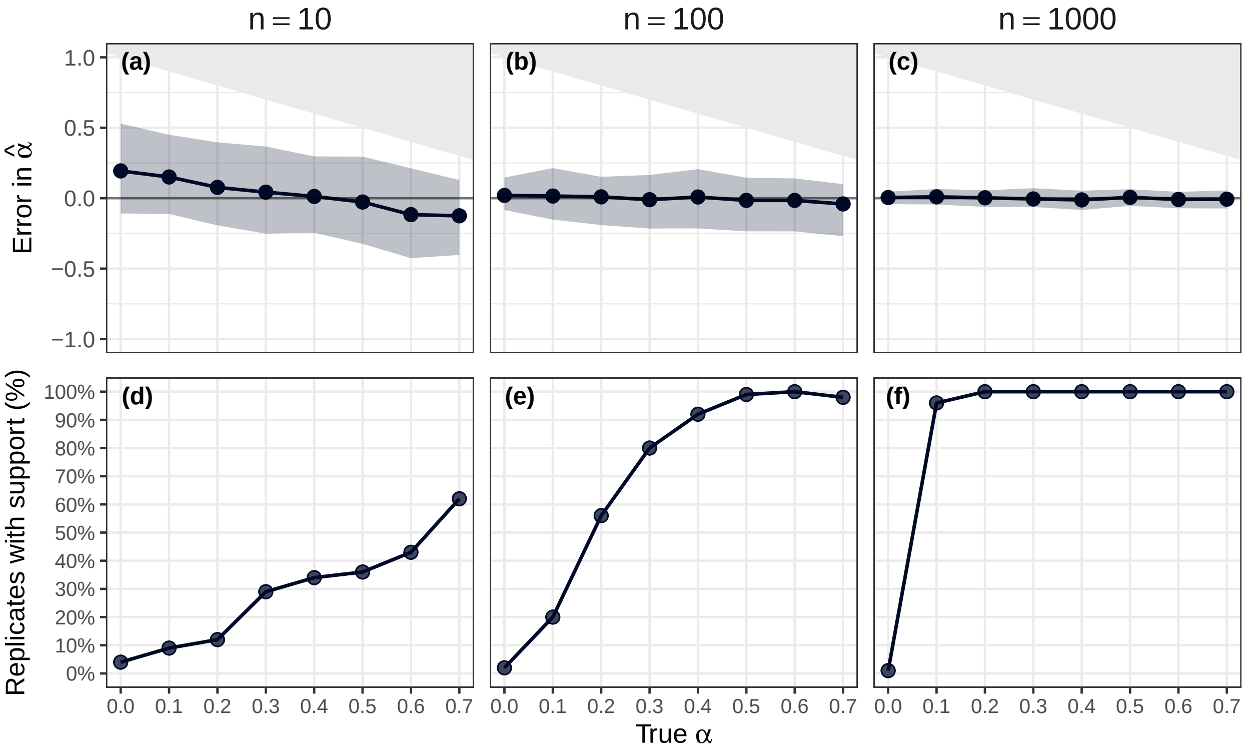

**Fig. S14. Error in the estimated values of $\hat{\alpha}$ and the percentage of replicates with support for symptom propagation, using an age-dependent methodology: $\mathbf{b = 4}$.** The top row shows the error in the estimates of $\alpha$. A positive error corresponds to the estimated value being an overestimate of the true value. The solid grey regions correspond to infeasible errors, which are present due to the restriction of $\hat{\alpha}$ being between 0 and 1. The points show the median errors and the lines show the interpolation between those points. The shaded area spans the 2.5th percentile to the 97.5th percentile (the 95% uncertainty interval). The bottom row gives the percentage of replicates (out of 1000) for which $\alpha = 0$ was not within the 95% confidence region (i.e. support for symptom propagation). Across panels, we vary the assessed sample sizes $N_{CT}$ (i.e. the number of data points used in each replicate). Each column corresponds to the number of contact tracing data points: **(a, d)** 10, **(b, e)** 100, **(c, f)** 1,000.

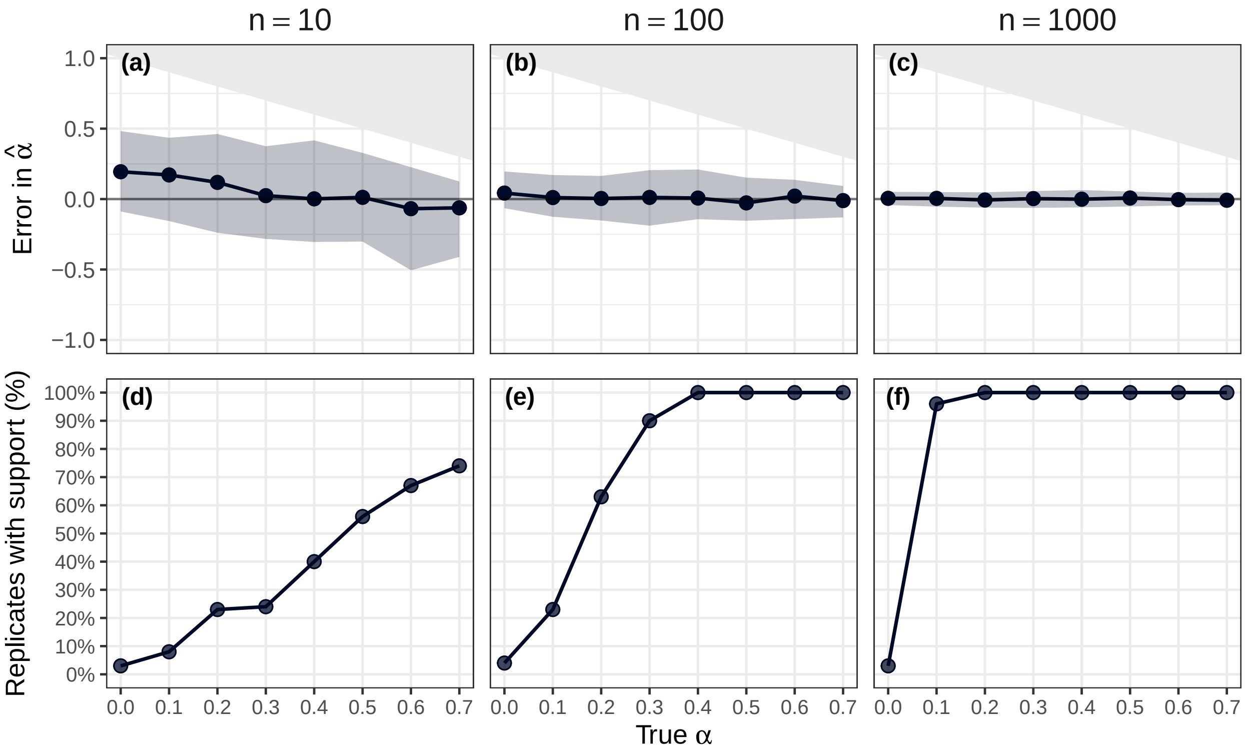

**Fig. S15. Error in the estimated values of $\hat{\alpha}$ and the percentage of replicates with support for symptom propagation, using an age-dependent methodology: $\mathbf{b = 1}$.** The top row shows the error in the estimates of $\alpha$. A positive error corresponds to the estimated value being an overestimate of the true value. The solid grey regions correspond to infeasible errors, which are present due to the restriction of $\hat{\alpha}$ being between 0 and 1. The points show the median errors and the lines show the interpolation between those points. The shaded area spans the 2.5th percentile to the 97.5th percentile (the 95% uncertainty interval). The bottom row gives the percentage of replicates (out of 1000) for which $\alpha = 0$ was not within the 95% confidence region (i.e. support for symptom propagation). Across panels, we vary the assessed sample sizes $N_{CT}$ (i.e. the number of data points used in each replicate). Each column corresponds to the number of contact tracing data points: **(a, d)** 10, **(b, e)** 100, **(c, f)** 1,000.

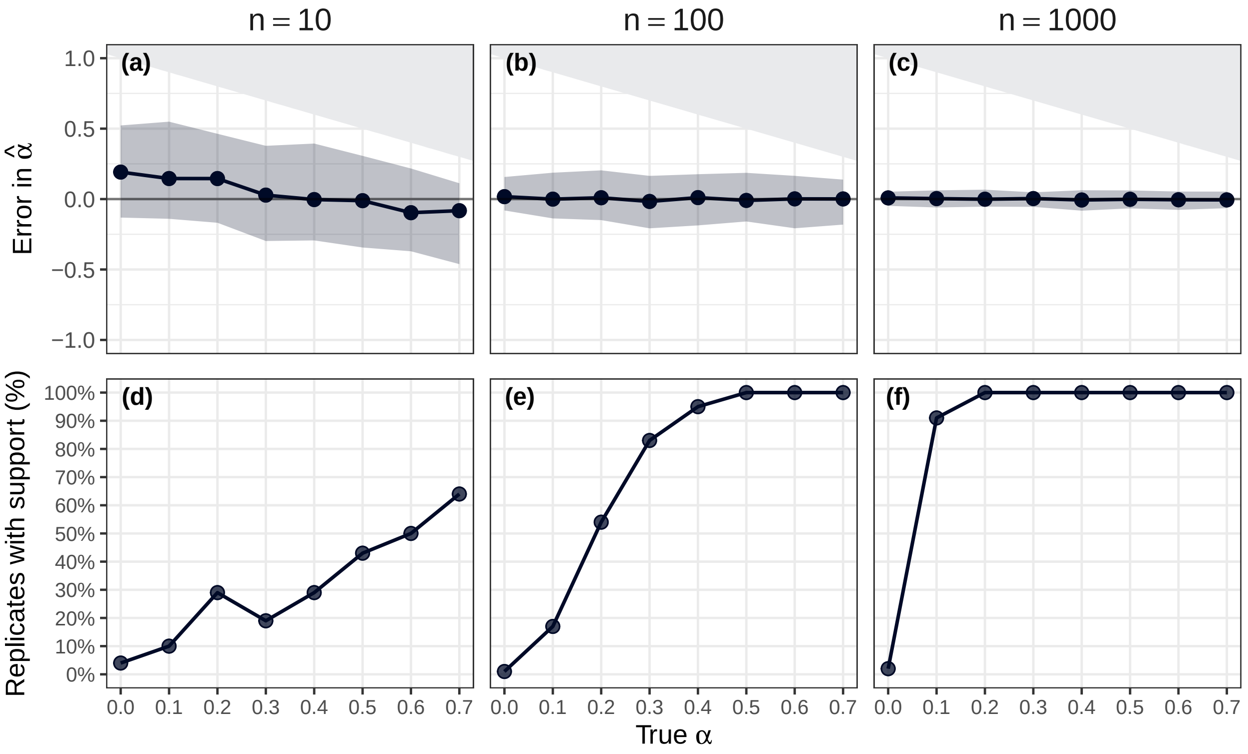

**Fig. S16. Error in the estimated values of $\hat{\alpha}$ and the percentage of replicates with support for symptom propagation, using an age-dependent methodology: $\mathbf{b = 0.5}$.** The top row shows the error in the estimates of $\alpha$. A positive error corresponds to the estimated value being an overestimate of the true value. The solid grey regions correspond to infeasible errors, which are present due to the restriction of $\hat{\alpha}$ being between 0 and 1. The points show the median errors and the lines show the interpolation between those points. The shaded area spans the 2.5th percentile to the 97.5th percentile (the 95% uncertainty interval). The bottom row gives the percentage of replicates (out of 1000) for which $\alpha = 0$ was not within the 95% confidence region (i.e. support for symptom propagation). Across panels, we vary the assessed sample sizes $N_{CT}$ (i.e. the number of data points used in each replicate). Each column corresponds to the number of contact tracing data points: **(a, d)** 10, **(b, e)** 100, **(c, f)** 1,000.

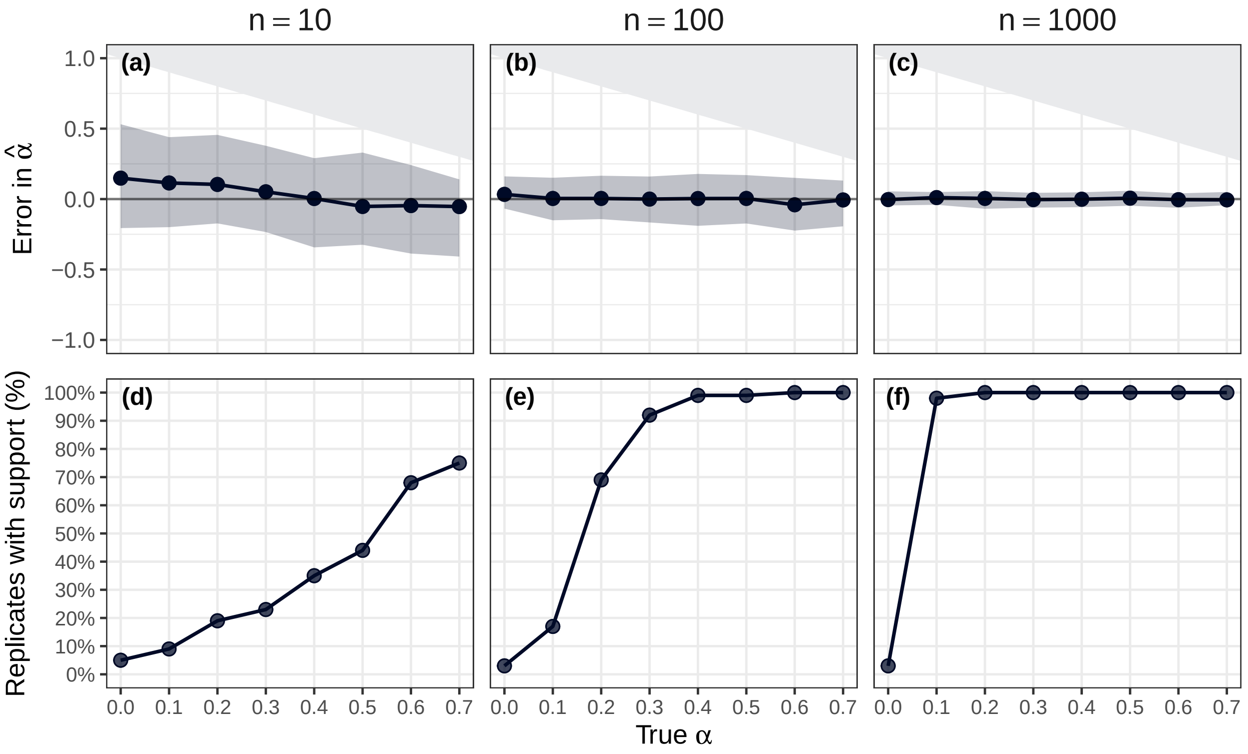

**Fig. S17. Error in the estimated values of $\hat{\alpha}$ and the percentage of replicates with support for symptom propagation, using an age-dependent methodology: $\beta = 0.5$.** The top row shows the error in the estimates of $\alpha$. A positive error corresponds to the estimated value being an overestimate of the true value. The solid grey regions correspond to infeasible errors, which are present due to the restriction of $\hat{\alpha}$ being between 0 and 1. The points show the median errors and the lines show the interpolation between those points. The shaded area spans the 2.5th percentile to the 97.5th percentile (the 95% uncertainty interval). The bottom row gives the percentage of replicates (out of 1000) for which $\alpha = 0$ was not within the 95% confidence region (i.e. support for symptom propagation). Across panels, we vary the assessed sample sizes $N_{CT}$ (i.e. the number of data points used in each replicate). Each column corresponds to the number of contact tracing data points: **(a, d)** 10, **(b, e)** 100, **(c, f)** 1,000.

###### S4.2.1 Moderate $\nu$ results

We additionally considered a scenario where symptom severity was less strongly age-dependent ($\nu = (0.2, 0.4, 0.6)$). We first provide a motivational plot showing the effect of underlying age-dependence in the data when an age-free methodology is used (Fig. S18). Then we provide the results for when the age-dependent methodology is used (Fig. S19).

Similarly to the main text $\nu$ values, we found that using an age-free methodology with age-dependent data led to an increase in the median error in $\alpha$ and an increase in the rate of finding support for symptom propagation when it does not exist, compared to when the data was age-free. However, the increase in the errors and rate of false positives was notably less than when the synthetic data was generated using more age-dependent values of $\nu$.

The results when the age-dependent methodology was used were virtually indistinguishable from the main text results (which used more age-dependent values of $\nu$).

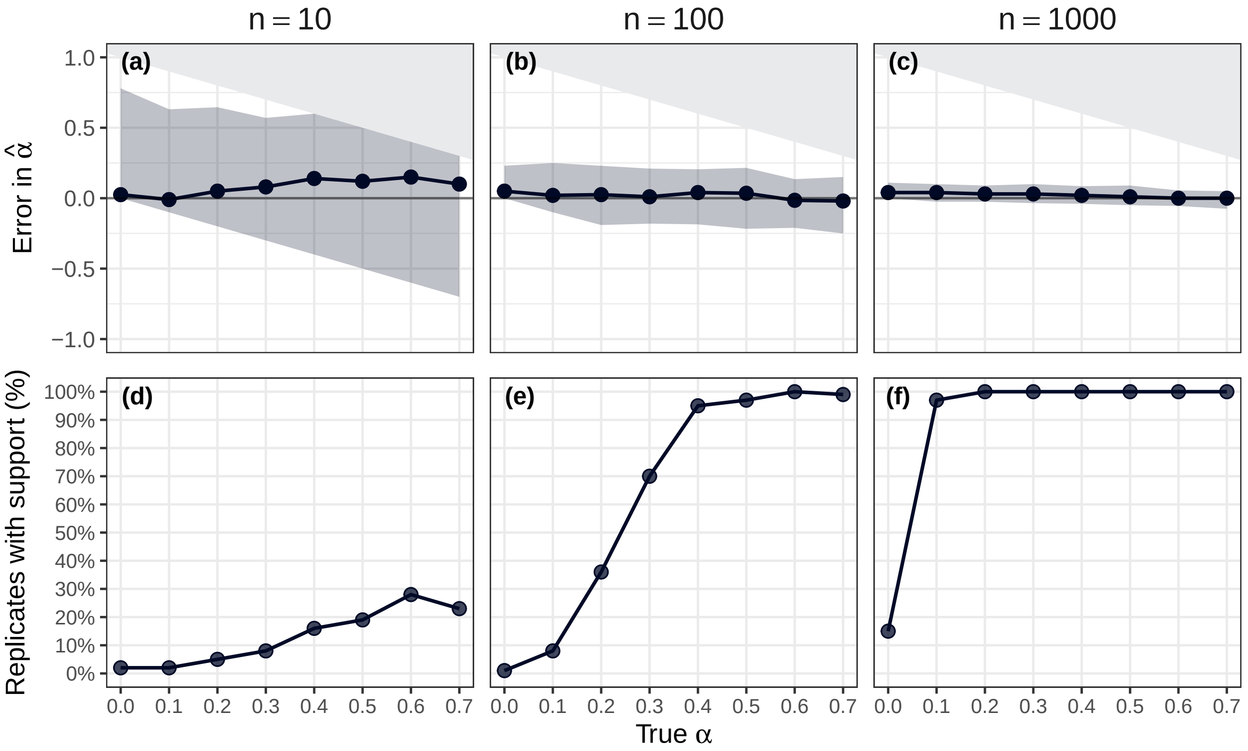

**Fig. S18. Error in the estimated values of $\hat{\alpha}$ and the percentage of replicates with support for symptom propagation, against the true values of $\alpha$: Age-free methodology and moderate $\nu$.** The top row shows the error in the estimates of $\alpha$. A positive error corresponds to the estimated value being an overestimate of the true value. The solid grey regions correspond to infeasible errors, which are present due to the restriction of $\hat{\alpha}$ being between 0 and 1. The points show the median errors and the lines show the interpolation between those points. The shaded area spans the 2.5th percentile to the 97.5th percentile (the 95% uncertainty interval). The bottom row gives the percentage of replicates (out of 1000) for which $\alpha = 0$ was not within the 95% confidence region (i.e. support for symptom propagation). Across panels, the assessed sample sizes $N_{CT}$ (i.e. the number of data points used in each replicate) vary. Each column corresponds to the number of contact tracing data points: **(a, d)** 10, **(b, e)** 100, **(c, f)** 1,000.

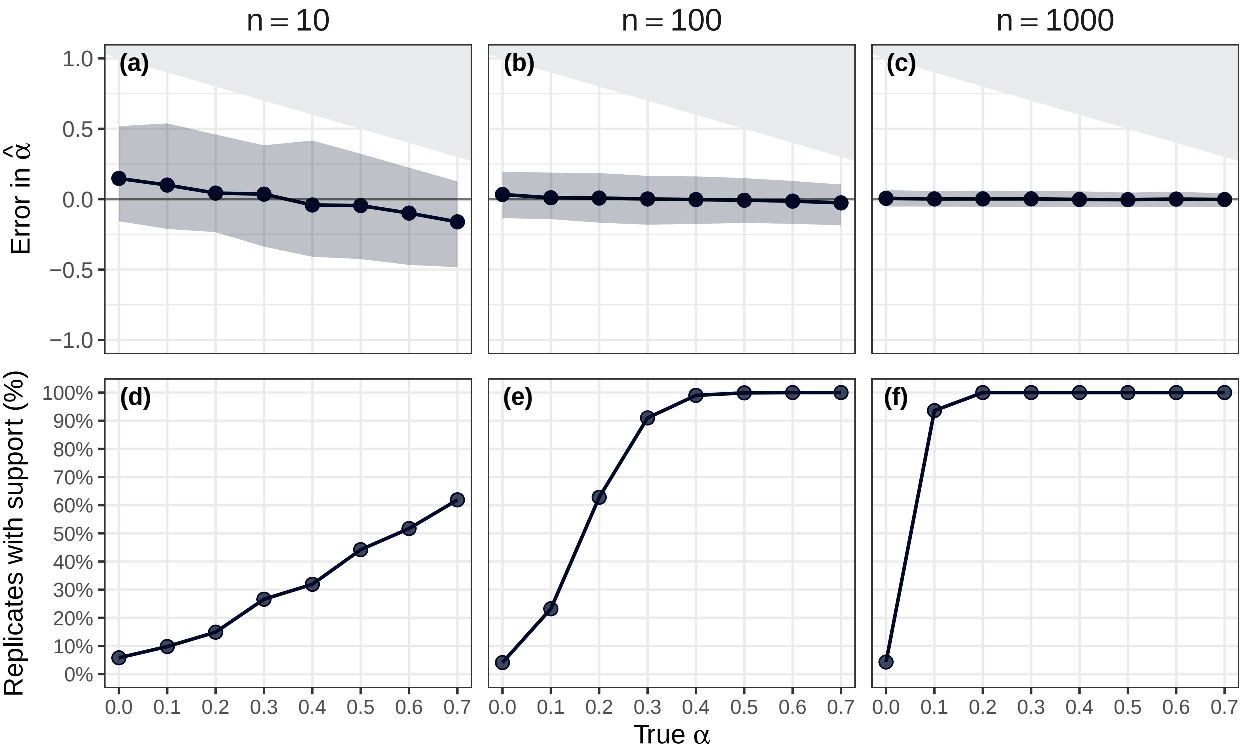

**Fig. S19. Error in the estimated values of $\hat{\alpha}$ and the percentage of replicates with support for symptom propagation, against the true values of $\alpha$: Moderate $\nu$.** The top row shows the error in the estimates of $\alpha$. A positive error corresponds to the estimated value being an overestimate of the true value. The solid grey regions correspond to infeasible errors, which are present due to the restriction of $\hat{\alpha}$ being between 0 and 1. The points show the median errors and the lines show the interpolation between those points. The shaded area spans the 2.5th percentile to the 97.5th percentile (the 95% uncertainty interval). The bottom row gives the percentage of replicates (out of 1000) for which $\alpha = 0$ was not within the 95% confidence region (i.e. support for symptom propagation). Across panels, the assessed sample sizes $N_{CT}$ (i.e. the number of data points used in each replicate) vary. Each column corresponds to the number of contact tracing data points: **(a, d)** 10, **(b, e)** 100, **(c, f)** 1,000.

#### S5 Additional real-world data results

##### S5.1 Trace plots for real-world data

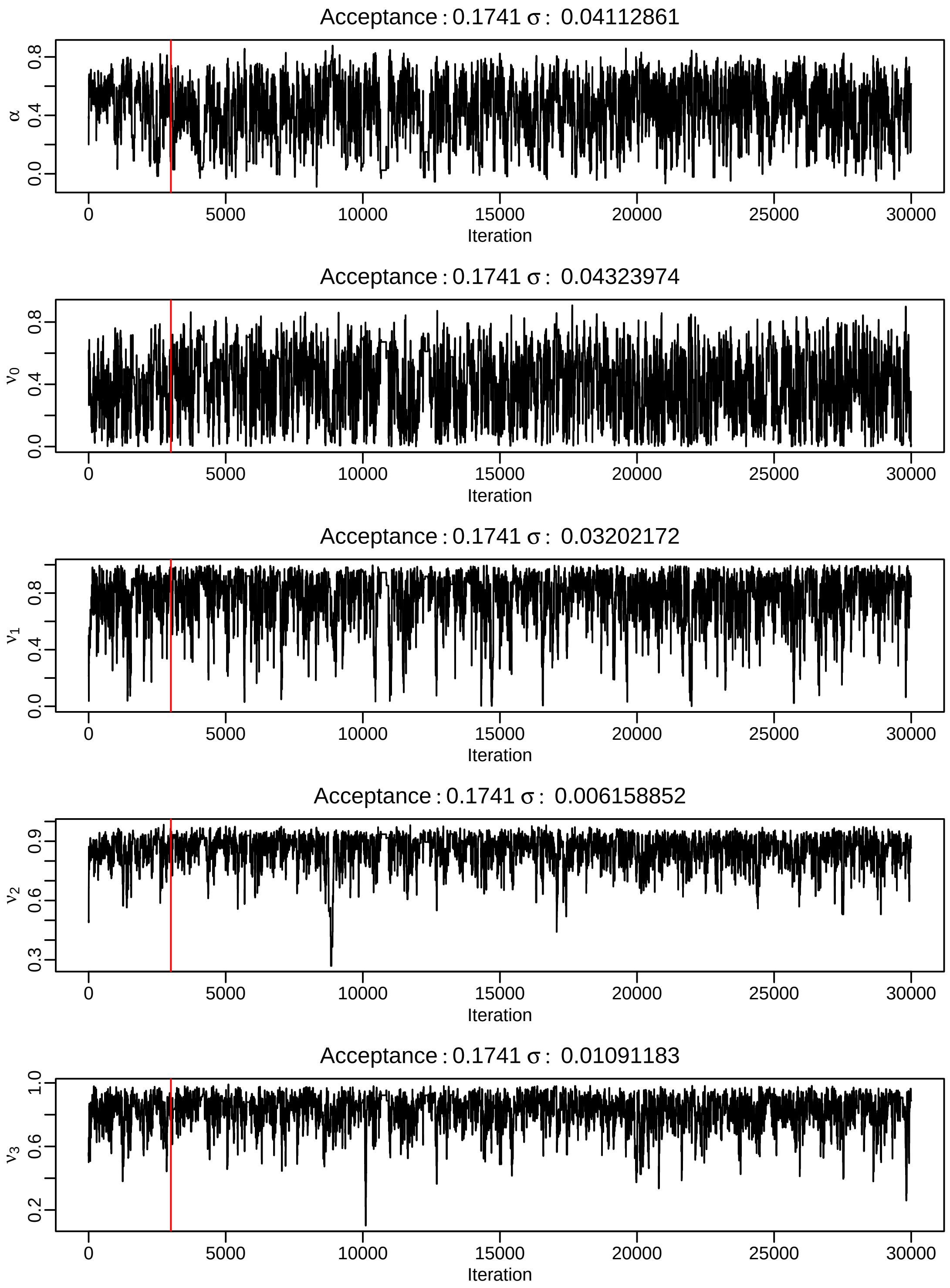

**Fig. S20. MCMC trace plots for parameter estimates from England households data.** Trace plots for $\alpha$ and age-dependent $\nu$ values. The red line indicates the end of the burn-in period (3,000 iterations).

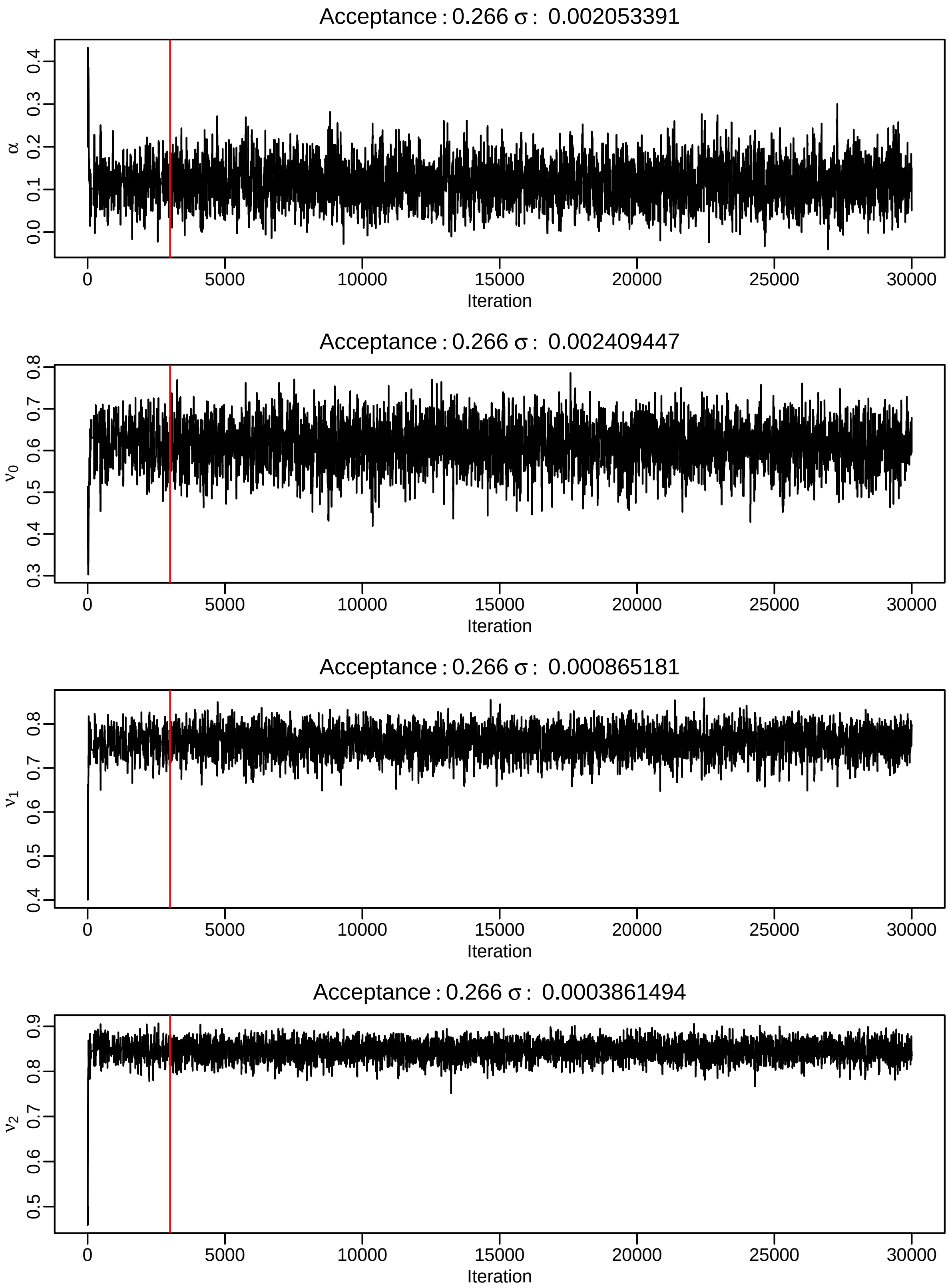

**Fig. S21. MCMC trace plots for parameter estimates from Israel households data.** Trace plots for $\alpha$ and age-dependent $\nu$ values. The red line indicates the end of the burn-in period (3,000 iterations).

#### S5.2 MCMC applied to age-free data

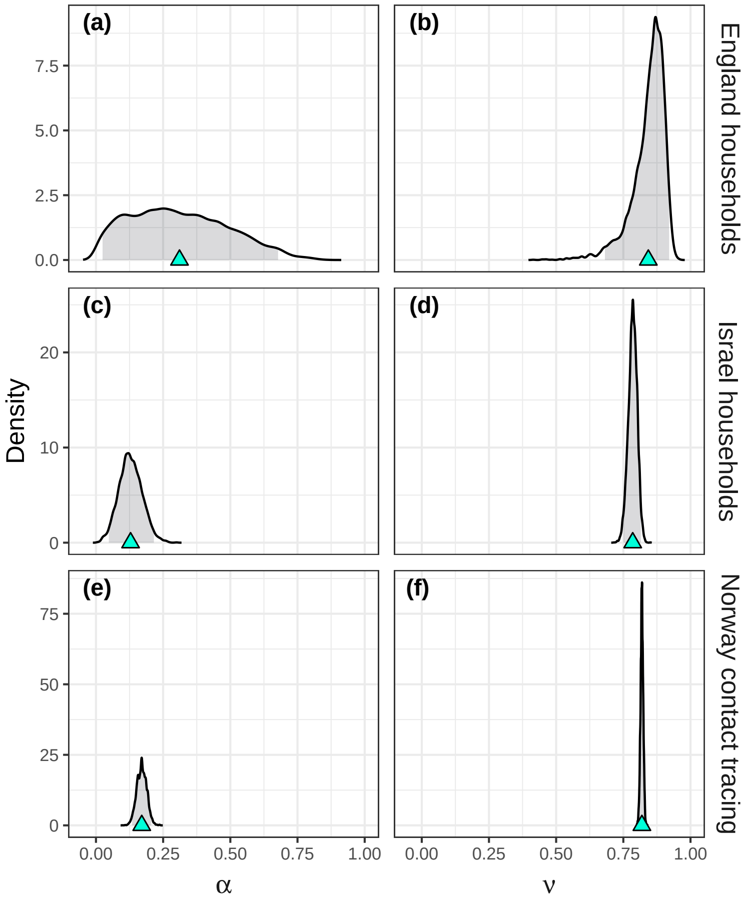

**Fig. S22. Posterior distributions for $\alpha$ and age-free $\nu$ values, for three real-world data sets.** Posterior distributions for $\alpha$ and a single age-free $\nu$ parameter for three real-world data sets: England households (top row), Israel households (middle row), and Norway contact tracing (bottom row). The cyan triangles indicate the posterior means and the grey shaded regions indicate the 95% credible intervals.
